# Visual art for the prevention of student mental health difficulties in schools: Mixed-methods analysis of staff surveys and interviews

**DOI:** 10.64898/2026.11.23.26368159

**Authors:** Bridget Ellis, Elizabeth Jestico, Dominic Treloar, Nikki McAuslan, Hester Careless, Emma Davies, Helen Bould, Judi Kidger, Naomi Warne

**Affiliations:** Centre for Academic Mental Health, Population Health Sciences, Bristol Medical School, University of Bristol, Bristol, UK; Butterfly Creative Therapy, UK; Independent researcher; St Cyres School, Cardiff, Wales; NIHR Bristol Biomedical Research Centre, University Hospitals Bristol and Weston NHS Foundation Trust and University of Bristol; MRC Epidemiology Unit, Population Health Sciences, Bristol Medical School, University of Bristol; Gloucestershire Health and Care NHS Foundation Trust, Gloucestershire; Centre for Public Health, Population Health Sciences, Bristol Medical School, University of Bristol, Bristol, UK

**Keywords:** Visual art, Mental health, School, Anxiety, Depression, Prevention, Intervention, Programme

## Abstract

**Background:** School prevention strategies are key to reducing the impact of youth mental health difficulties, such as depression and anxiety. Visual art programmes may be beneficial for prevention but evidence is limited. To better understand the potential for visual arts-based prevention programmes in schools, we explored staff experiences of using visual art for mental health and views on what is needed to facilitate successful programmes.

**Methods:** We used a convergent mixed-methods approach, combining online surveys (n=77) and in-depth interviews (n=27) from art teachers and staff supporting student mental health in UK secondary schools. Complementary survey and interview questions covered current use of visual art for student mental health, what a visual art prevention programme might look like, and potential concerns. We conducted descriptive statistics for quantitative survey data and reflexive thematic analysis for qualitative survey and interview data.

**Results:** Visual arts were seen as important for student mental health and commonly used: 65.8% of staff used visual arts to support student mental health in the last year, and 31.6% did so weekly, despite most participants reporting no relevant training. Theme 1, ‘Art is a sanctuary in the school day’, considered art as intrinsically linked to mental health, with art freedom conferring particular benefits. Theme 2, ‘Resources in schools are stretched’, represented the pressure of school environments, limited resources and need for prevention. Theme 3, ‘Integration, flexibility and relationships as factors for success’, highlighted integrating programmes into school practice, enabling flexibility and choice, and building positive relationships between students, school staff and external providers are key for success.

**Conclusions:** Despite implementation challenges, visual arts-based prevention approaches were viewed positively. We recommend school mental health arts programmes should promote school ownership, include creative freedom, be flexible to schools’ and students’ needs, and be supported by both internal school staff and external providers.

## Background

Adolescent anxiety and depression are common and are associated with significant impairment, long-term mental health difficulties and education and employment problems (1–5). Peak age at onset for social anxiety disorder, panic disorder, generalised anxiety disorder, and depressive disorders is in adolescence (6). Primary prevention programmes are needed to reduce the burden and impact of mental illness in young people (7). The World Health Organization (2020) advocates prioritising prevention strategies for adolescent mental health conditions and integrated approaches towards this, including educational and health care services (8). Schools offer one way to implement prevention programmes at scale. There is evidence supporting school-based prevention programmes for primary prevention generally (7) and some evidence supporting programmes preventing anxiety and depression (9,10). However, many programmes lack longevity (10) and prevention approaches that seem promising in small-scale trials often fail to replicate positive findings in large, well-powered studies (11,12). Such programmes often focus on using treatment approaches (e.g. cognitive behavioural therapy, mindfulness) in a prevention context (9), whereas a focus on modifiable risk factors for anxiety and depression may be more effective for those who have not yet developed mental health difficulties depression (11). For instance, a whole-school approach using restorative practice and social and emotional learning to target bullying was effective at reducing both bullying and mental health difficulties three years later (13).

There is growing evidence connecting the arts (14–17) and art therapy (18,19) to positive mental health and wellbeing outcomes in young people, suggesting arts-based interventions may target risk factors for anxiety and depression. The multimodal nature of arts means that engagement can affect health through diverse psychological, biological, social and behavioural mechanisms (20,21). Importantly, for children and young people, visual arts (for instance, painting, drawing, crafting, design, photography, sculpture, and textiles (21)) offer an opportunity to communicate without the need for words, a useful mechanism for young people who find it difficult to label emotions and describe experiences. Furthermore, engaging in art and art therapy can increase effective emotion regulation (22–24), improve self-esteem (14,25–28) and reduce stress (29,30). These factors may, in turn, buffer against development of anxiety and depression (31–35). Therefore, targeting these protective factors using engagement with visual arts may be a useful prevention strategy. There is preliminary evidence that students with emotional/behavioural problems receiving preventative art therapy in schools from art therapists show improvements in mental health (36,37), but little is known about using arts as a universal prevention strategy in schools or prevention strategies not delivered by art therapists.

School buy in, support from multiple levels of the school community, and competency of teachers can help school programmes achieve longevity, uptake and success (38–41). However, there is some evidence that school staff delivery may be less effective than external provider delivery (10,42), potentially due to low levels of teacher acceptability and confidence in delivering interventions, alongside inadequate time, training and support (43). Arts-based programmes, in particular, may face implementation challenges given the evidence that the arts are not valued in the English state education system (44). An in-depth understanding of the experiences and perspectives of staff working in and with schools is needed to determine how to make an art and wellbeing prevention programme in schools successful and sustainable. These staff groups are key for exploring the current cultural attitudes towards visual arts and mental health, and navigating the barriers to implementing arts programmes within schools.

### The current study

The aims of this study were to (1) explore current experiences of using visual art to promote student mental health in UK secondary schools and (2) better understand what would be needed to facilitate a school-based art and wellbeing programme to help prevent anxiety and depression. We chose a mixed methods design to enable both breadth and depth in understanding staff’s experiences and perspectives. We collected quantitative and qualitative data via an online survey and interviews with staff across the UK from staff who would be closely involved in the organisation or delivery of an art and wellbeing prevention programme.

## Methods

### Study design

We used a convergent mixed methods design to collect quantitative data (survey) and qualitative (survey and interview) data during one phase of data collection. Survey and interview questions were designed to collect parallel data using similar questions, with the survey enabling views from a wider range of staff and interviews providing more depth and contextualised exploration of the research topic. We pre-registered the Study Protocol prior to data collection on the Open Science Framework: https://osf.io/erq5a/ and used Good Reporting of A Mixed Methods Study (GRAMMS) guidelines (45) (S1 Table).

### Participants and recruitment

We recruited participants who were over 18 years old and worked in a UK secondary school as either an art teacher specialising in visual or fine arts, or in a role supporting student mental health. This second broad category included staff based within schools (for example, mental health leads, teaching assistants and Special Educational Needs Coordinators (SENCOs)) and external professionals (for example, creative arts therapists and Educational Mental Health Practitioners (EMHPs)). Participants were excluded if they did not speak fluent English, or they were public contributors involved in designing the study materials.

Recruitment for surveys and interviews took place between June 2023 and January 2024 by (1) utilising existing networks and contacts to share study information, (2) directly contacting UK schools and external organisations with links to schools to share information with staff, (3) sharing through established school research networks and (4) sharing on social media. We also asked participants to share study information with other potentially eligible individuals.

Participant recruitment is presented in S1 Figure. Potential participants initially registered their interest by completing a short survey with demographic information, occupation, and preference to take part in the survey and/or interview. Verification of eligibility and genuine participant status was manually conducted by checking staff lists on school websites and work email addresses. All verified and eligible individuals expressing an interest in survey participation were sent a link to complete the online survey, and a reminder email after two weeks if the survey had not been completed. We carried out purposive sampling to ensure a diverse range of views across interview participants. Verified and eligible individuals were invited to book an interview time if they had a job role or demographic characteristics (age, gender, ethnicity, region of work) not yet included in the interview sample, with a reminder email after two weeks if they had not booked an interview time. Participants were remunerated with a £10 voucher for completing the survey and a £25 voucher for completing an interview.

A total of 79 participants completed the online survey, of whom 28 individuals were also interviewed. The research team were mindful of fraudulent participants throughout data collection, which resulted in data from two participants being excluded, one due to substantial discrepancies between registration of interest form and survey responses (survey only) and the other as an interview participant presenting with vague responses, not answering questions on topic, and avoiding sharing information about work (survey and interview).

The final sample included 77 survey participants and 27 interview participants. Professional backgrounds varied (Table 1 and S2 Table). Most were female (93%), white British (survey 84%, interview 74%), had worked in education for 10 years or more (55% survey, 54% interview), and had worked in England (92% survey, 85% interview). Characteristics of those interviewed were similar to those participating in the survey (Table 1).

**Table 1.**
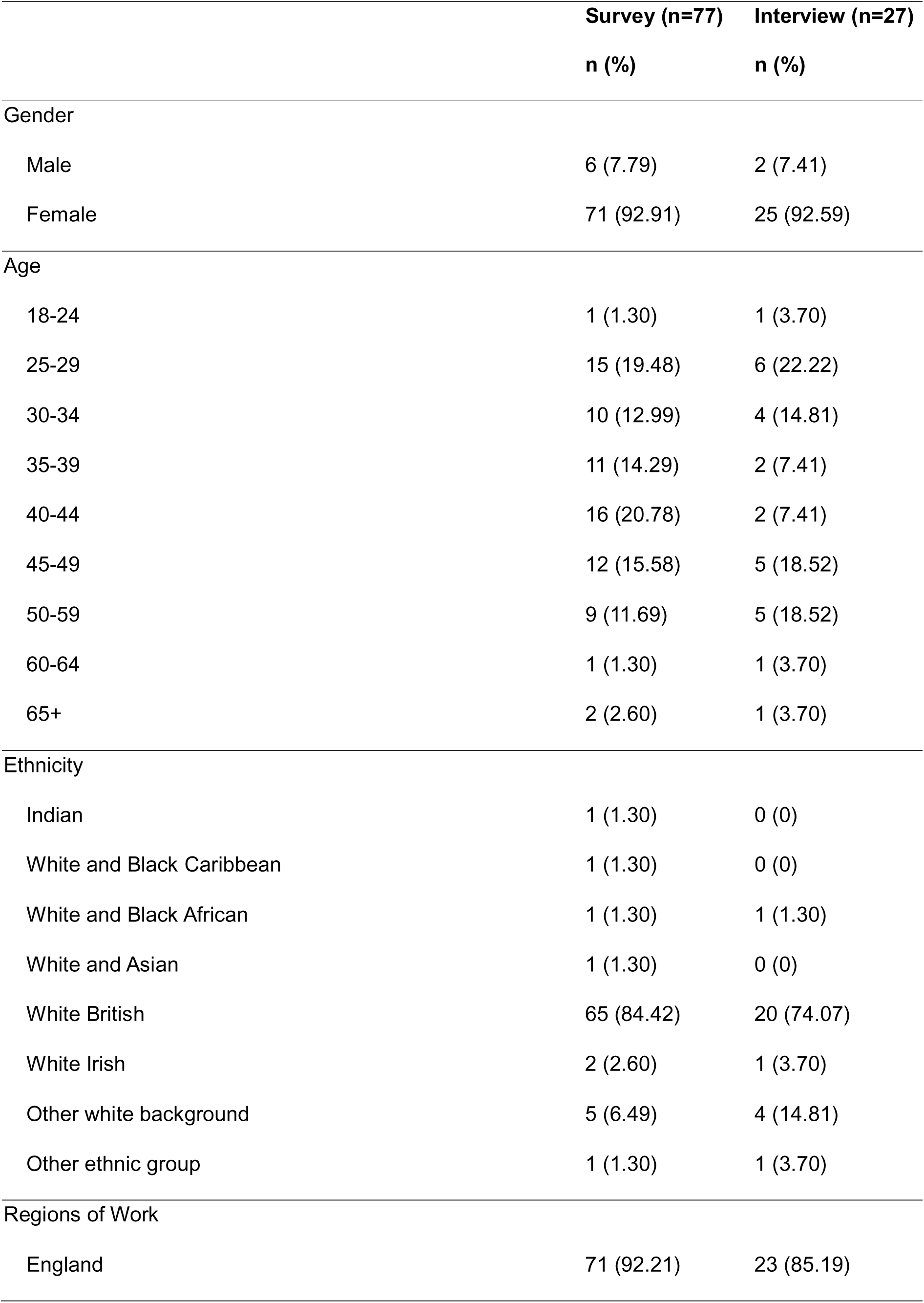
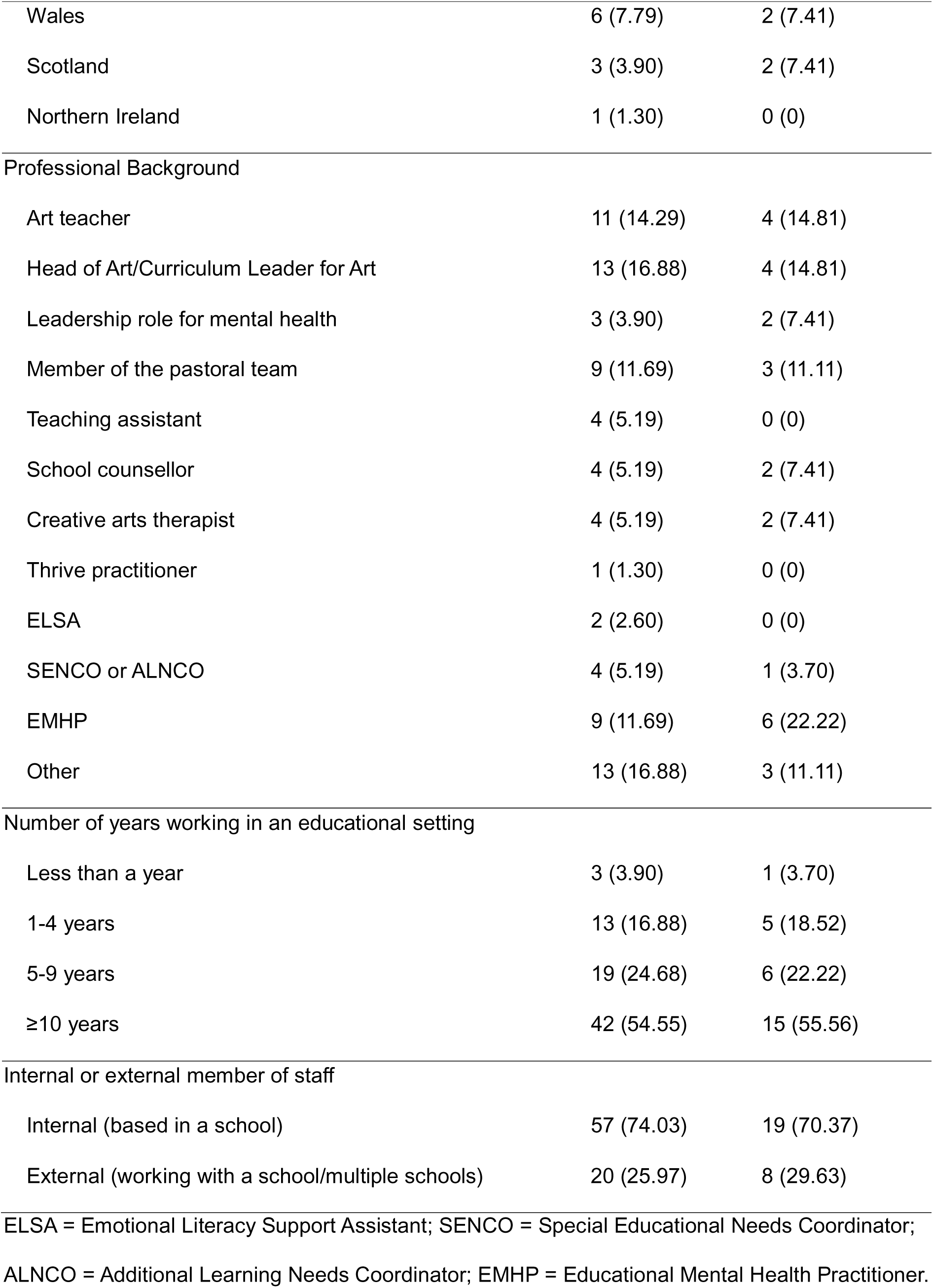
Demographic information for survey (n=77) and interview (n=27) participants.

|  | **Survey (n=77)**<br>**n (%)** | **Interview (n=27)**<br>**n (%)** |
|---|---|---|
| Gender |  |  |
| Male | 6 (7.79) | 2 (7.41) |
| Female | 71 (92.91) | 25 (92.59) |
| Age |  |  |
| 18-24 | 1 (1.30) | 1 (3.70) |
| 25-29 | 15 (19.48) | 6 (22.22) |
| 30-34 | 10 (12.99) | 4 (14.81) |
| 35-39 | 11 (14.29) | 2 (7.41) |
| 40-44 | 16 (20.78) | 2 (7.41) |
| 45-49 | 12 (15.58) | 5 (18.52) |
| 50-59 | 9 (11.69) | 5 (18.52) |
| 60-64 | 1 (1.30) | 1 (3.70) |
| 65+ | 2 (2.60) | 1 (3.70) |
| Ethnicity |  |  |
| Indian | 1 (1.30) | 0 (0) |
| White and Black Caribbean | 1 (1.30) | 0 (0) |
| White and Black African | 1 (1.30) | 1 (1.30) |
| White and Asian | 1 (1.30) | 0 (0) |
| White British | 65 (84.42) | 20 (74.07) |
| White Irish | 2 (2.60) | 1 (3.70) |
| Other white background | 5 (6.49) | 4 (14.81) |
| Other ethnic group | 1 (1.30) | 1 (3.70) |
| Regions of Work |  |  |
| England | 71 (92.21) | 23 (85.19) |
| Wales | 6 (7.79) | 2 (7.41) |
| Scotland | 3 (3.90) | 2 (7.41) |
| Northern Ireland | 1 (1.30) | 0 (0) |
| Professional Background |  |  |
| Art teacher | 11 (14.29) | 4 (14.81) |
| Head of Art/Curriculum Leader for Art | 13 (16.88) | 4 (14.81) |
| Leadership role for mental health | 3 (3.90) | 2 (7.41) |
| Member of the pastoral team | 9 (11.69) | 3 (11.11) |
| Teaching assistant | 4 (5.19) | 0 (0) |
| School counsellor | 4 (5.19) | 2 (7.41) |
| Creative arts therapist | 4 (5.19) | 2 (7.41) |
| Thrive practitioner | 1 (1.30) | 0 (0) |
| ELSA | 2 (2.60) | 0 (0) |
| SENCO or ALNCO | 4 (5.19) | 1 (3.70) |
| EMHP | 9 (11.69) | 6 (22.22) |
| Other | 13 (16.88) | 3 (11.11) |
| Number of years working in an educational setting |  |  |
| Less than a year | 3 (3.90) | 1 (3.70) |
| 1-4 years | 13 (16.88) | 5 (18.52) |
| 5-9 years | 19 (24.68) | 6 (22.22) |
| ≥10 years | 42 (54.55) | 15 (55.56) |
| Internal or external member of staff |  |  |
| Internal (based in a school) | 57 (74.03) | 19 (70.37) |
| External (working with a school/multiple schools) | 20 (25.97) | 8 (29.63) |

ELSA = Emotional Literacy Support Assistant; SENCO = Special Educational Needs Coordinator; ALNCO = Additional Learning Needs Coordinator; EMHP = Educational Mental Health Practitioner.

## Data collection

### Survey

Participants completed an online survey covering questions about (1) their demographic characteristics and professional background, (2) current experiences around arts and mental health in schools (frequency of using arts in their job, perceived importance of visual arts for student mental health, current use of visual art to support student mental health, current experiences of supporting student mental health and training), (3) what an art and wellbeing programme should look like (who should support/deliver the programme, group size, time of day, resources) and (4) any concerns about an art and wellbeing programme. Questions were a mixture of forced-choice, unrestricted choice and open-ended questions with free text boxes for participants to express their opinions. There was open text box at the end of the survey for participants to provide additional information. Full questions are reported in S1 File. Open text questions broadly aligned with areas and questions on the interview topic guide (see below).

### Interview

Three researchers (BE, HC and NW) conducted semi-structured interviews over phone or video call (Teams/Zoom). The interviews were conducted via the flexible use of a topic guide (S2 File) and probing to gather more detail around areas of interest. We used similar questions and topics to those used in the survey, including (1) visual art in the context of mental health (potential links, current experiences of using visual art to support student mental health), (2) what an art and wellbeing programme should look like (who should support/deliver the programme, group size, time of day, sustainability, engaging students, implementation) and (3) any concerns about an art and wellbeing programme (including school mental health safeguarding and policies). Participants were given a list of starting questions before the interview as this was highlighted by our public contributors as a way to facilitate more in-depth interview discussions. As all interview participants completed the survey first, we used their responses to guide the interview and identify areas where more in-depth discussion would be beneficial.

### Data analysis

Separate quantitative and qualitative data analyses were conducted in parallel before being integrated to enable one overall interpretation in the Discussion. Priority was given to the qualitative results which provided depth and context to the quantitative results.

### Quantitative data

Descriptive statistics were performed on the quantitative survey using Stata version 18 (46) by NW. Percentages were calculated for fixed-choice and unrestricted choice answers for the total sample, as well as separately for internal staff working within a school and external staff working with a school or multiple schools (variable derived from survey and interview responses). We chose not to use inferential statistics to examine differences between groups due to small sample sizes and lack of power.

### Qualitative data

Given the alignment of open text survey questions and interview questions, we chose to pool and analyse data from open-text survey responses and interview transcripts as one dataset in NVivo. BE led Reflexive Thematic Analysis (47,48) of the data, with regular discussions with NW and JK. By using Reflexive Thematic Analysis, we acknowledge that codes and themes will reflect the pre-existing knowledge, experiences and social position of the researchers and this reflexivity is integral to our interpretation of the data. Our reflexive positioning is presented in S3 Table.

Throughout the data analysis, BE read both hard copies of the transcripts and electronic copies on NVivo to prompt new insights and reflections. BE listened to recordings and read transcripts for data familiarisation, making notes of initial thoughts and reflections. From this a process of systematic coding began, identifying patterns of meaning within the data. Once the codes fully captured all the data, initial themes were generated and discussed in meetings with NW and JK. The themes were then developed, and through a process of presenting coding, themes and extracts from the transcripts, the themes were refined and reviewed collaboratively with members of the wider research team. This was an iterative and recursive process, going between the themes, the data and reflexive notes throughout.

### Integration

Quantitative and qualitative results were integrated during the interpretation phase by NW and further reviewed, edited and interpreted by the wider team. We synthesised both data types to provide recommendations for developing school-based arts programmes.

### Patient and public involvement (PPI)

We consulted multiple key informants when developing the project, including young people (n=6, aged 11-16 years old) in a PPI session and adult stakeholders (an art therapist, art teachers (n=3), a child and adolescent psychiatrist, and an educational mental health practitioner) via one-to-one discussions. These discussions informed key topics to include in the current and subsequent studies.

Alongside input from the research team, we received feedback on the study materials from a secondary school Head of Art, a teaching assistant, and a teacher who is member of their school’s pastoral team. Changes implemented from this feedback included ensuring terminology was consistent with practice, clarifying specific questions around the arts and visual arts, simplifying the participant information sheets and adding questions to better understand the school/work context.

### Ethics approval and consent to participate

Ethic approval was obtained from University of Bristol Faculty of Health Sciences Research Ethics Committee (Reference 13484) and Bristol City Council Research Governance (Reference 2023-005). All participants provided written informed consent.

## Results

### Quantitative survey results

Full quantitative results from the survey are presented in Table 2. Participants commonly used the arts in their work, with 48.05% reporting using the arts “a lot” and 42.86% using the *visual* arts “a lot”. Most respondents (88.31%) felt visual art was important (53.25%) or very important (35.06%) for supporting student mental health. This translated to art and creative activities being used in the context of mental health: almost two thirds of staff (65.79%) reported using visual arts to promote or support student mental health in the past year, with almost a third of staff (31.58%) reporting use at least once a week. This was often informal or something for which staff had not received training: 72.73% report no student mental health training incorporating the arts and 89.47% report no teacher mental health training incorporating the arts.

**Table 2.** Survey responses on art and mental health, current mental health in schools and references for an art and wellbeing programme (n=77)

| **Survey item and response** | **Total survey sample (n=77)** | **Internal staff<sup>a</sup> (n=57)** | **External staff<sup>b</sup> (n=20)** |
|---|---|---|---|
|  | **n (%)** | **n (%)** | **n (%)** |
| **Current use of the arts for work (e.g. creative writing, experiencing music, creating images)** |  |  |  |
| None | 4 (5.19) | 4 (7.02) | 0 (0.00) |
| A little | 20 (25.97) | 13 (22.81) | 7 (35.00) |
| Some | 16 (20.78) | 10 (17.54) | 6 (30.00) |
| A lot | 37 (48.05) | 30 (52.63) | 7 (35.00) |
| **Current use of visual arts for work (e.g. creating images or sculptures, drawing, painting)** |  |  |  |
| None | 9 (11.69) | 8 (14.04) | 1 (5.00) |
| A little | 22 (28.57) | 13 (22.81) | 9 (45.00) |
| Some | 13 (16.88) | 8 (14.04) | 5 (25.00) |
| A lot | 33 (42.86) | 28 (49.12) | 5 (25.00) |
| **Importance of creating visual art for supporting student mental health** |  |  |  |
| Very unimportant | 3 (3.90) | 3 (5.26) | 0 (0.00) |
| Unimportant | 0 (0) | 0 (0.00) | 0 (0.00) |
| Neutral | 6 (7.79) | 4 (7.02) | 2 (10.00) |
| Important | 41 (53.25) | 30 (52.63) | 11 (55.00) |
| Very important | 27 (35.06) | 20 (35.09) | 7 (35.00) |
| **Frequency of using visual art as a tool to promote or support student mental health in the past year<sup>a</sup>** |  |  |  |
| Never | 26 (34.21) | 25 (43.86) | 1 (5.26) |
| Less than once a month | 12 (15.79) | 7 (12.28) | 5 (26.32) |
| Once a month | 8 (10.53) | 6 (10.53) | 2 (10.53) |
| 2-3 times a month | 6 (7.89) | 6 (10.53) | 0 (0.00) |
| Once a week | 7 (9.21) | 6 (10.53) | 1 (5.26) |
| 2-4 times a week | 7 (9.21) | 3 (5.26) | 4 (21.05) |
| Once a day | 0 (0) | 0 (0.00) | 0 (0.00) |
| Multiple times a day | 10 (13.16) | 4 (7.02) | 6 (31.58) |
| **Received training in supporting student mental health that incorporated creativity or use of the arts** |  |  |  |
| No | 56 (72.73) | 50 (87.72) | 6 (30.00) |
| Yes | 21 (27.27) | 7 (12.28) | 14 (70.00) |
| **Received training in supporting teacher/staff mental health that incorporated creativity or use of the arts<sup>c</sup>** |  |  |  |
| No | 68 (89.47) | 54 (94.74) | 14 (73.68) |
| Yes | 8 (10.53) | 3 (5.26) | 5 (26.32) |
| **Staff who should be involved in *delivering* an art-based programme to help prevent student anxiety and depression (multiple answers possible)** |  |  |  |
| School mental health support staff | 62 (80.52) | 45 (78.95) | 17 (85.00) |
| Mental health leaders | 42 (54.55) | 31 (54.39) | 11 (55.00) |
| Head of Art/Curriculum Leaders for Art | 33 (42.86) | 28 (49.12) | 5 (25.00) |
| Pastoral team | 42 (54.55) | 31 (54.39) | 11 (55.00) |
| Art teachers | 52 (67.53) | 41 (71.93) | 11 (55.00) |
| Non-art teachers | 19 (24.68) | 14 (24.56) | 5 (25.00) |
| Teaching support staff | 37 (48.05) | 29 (50.88) | 8 (40.00) |
| External providers/specialists | 37 (48.05) | 24 (42.11) | 13 (65.00) |
| Other | 3 (3.90) | 1 (1.75) | 2 (10.00) |
| **Staff who should be involved in *supporting* an art-based programme to help prevent student anxiety and depression (multiple answers possible)** |  |  |  |
| School mental health support staff | 70 (90.91) | 51 (89.47) | 19 (95.00) |
| Mental health leaders | 67 (87.01) | 48 (84.21) | 19 (95.00) |
| Head of Art/Curriculum Leaders for Art | 50 (64.94) | 39 (68.42) | 11 (55.00) |
| Pastoral team | 55 (71.43) | 41 (71.93) | 14 (70.00) |
| Art teachers | 62 (80.52) | 45 (78.95) | 17 (85.00) |
| Non-art teachers | 31 (40.26) | 19 (33.33) | 12 (60.00) |
| Teaching support staff | 51 (66.23) | 35 (61.40) | 16 (80.00) |
| External providers/specialists | 47 (61.04) | 31 (54.39) | 16 (80.00) |
| Other | 5 (6.49) | 2 (3.51) | 3 (15.00) |
| **Frequency of mental health and wellbeing conversations with students in the past year** |  |  |  |
| Never | 1 (1.30) | 1 (1.75) | 0 (0.00) |
| Less than once a month | 4 (5.19) | 4 (7.02) | 0 (0.00) |
| Once a month | 3 (3.90) | 3 (5.26) | 0 (0.00) |
| 2-3 times a month | 9 (11.69) | 7 (12.28) | 2 (10.00) |
| Once a week | 7 (9.09) | 5 (8.77) | 2 (10.00) |
| 2-4 times a week | 15 (19.48) | 15 (26.32) | 0 (0.00) |
| Once a day | 8 (10.39) | 7 (12.28) | 1 (5.00) |
| Multiple times a day | 30 (38.96) | 15 (26.32) | 15 (75.00) |
| **Confidence talking to students about mental health problems** |  |  |  |
| Not at all confident | 2 (2.60) | 2 (3.51) | 0 (0.00) |
| Somewhat confident | 15 (19.48) | 15 (26.32) | 0 (0.00) |
| Moderately confident | 27 (35.06) | 21 (36.84) | 6 (30.00) |
| Extremely confident | 33 (42.86) | 19 (33.33) | 14 (70.00) |
| **Days of training to support student mental health in the past three years** |  |  |  |
| None | 12 (15.58) | 11 (19.30) | 1 (5.00) |
| Less than a day | 9 (11.69) | 9 (15.79) | 0 (0.00) |
| 1 day | 6 (7.79) | 6 (10.53) | 0 (0.00) |
| 2 days | 4 (5.19) | 4 (7.02) | 0 (0.00) |
| 3 days | 12 (15.58) | 11 (19.30) | 1 (5.00) |
| 4 days | 5 (6.49) | 5 (8.77) | 0 (0.00) |
| 5 days | 4 (5.19) | 2 (3.51) | 2 (10.00) |
| 6+days | 25 (32.47) | 9 (15.79) | 16 (80.00) |
| **Want mental health training to support students** |  |  |  |
| No | 31 (40.26) | 24 (42.11) | 7 (35.00) |
| Yes | 46 (59.74) | 33 (57.89) | 13 (65.00) |
| **Size of group for art programme (forced choice)<sup>d</sup>** |  |  |  |
| Whole classes | 9 (11.84) | 7 (12.50) | 2 (10.00) |
| Smaller groups | 67 (88.16) | 49 (87.50) | 18 (90.00) |
| **Best time of day for an art programme delivery (multiple answers)** |  |  |  |
| PSHE or health lessons | 39 (50.65) | 26 (45.61) | 13 (65.00) |
| Art lessons | 19 (24.68) | 17 (29.82) | 2 (10.00) |
| Tutor time | 29 (37.66) | 24 (42.11) | 5 (25.00) |
| Half day activity | 37 (48.05) | 26 (45.61) | 11 (55.00) |
| Whole day activity | 33 (42.86) | 25 (43.86) | 8 (40.00) |
| Extracurricular club after school | 32 (41.56) | 21 (36.84) | 11 (55.00) |
| Extracurricular club during lunch | 30 (38.96) | 18 (31.58) | 12 (60.00) |
| Other | 16 (20.78) | 8 (14.04) | 8 (40.00) |

<sup>a</sup> Internal staff = staff predominantly based and integrated within one school setting
<sup>b</sup> External staff = staff working with a school or multiple schools but not based within the school setting
<sup>c</sup> total sample n=76 and internal staff n = 56 due to missing data
<sup>d</sup> total sample n=76 and external staff n = 19 due to missing data

Quantitative results suggest mixed opinions on who would be needed to facilitate an art and wellbeing programme in schools. Less than half of survey respondents (48.05%) thought external providers should deliver an art-based mental health prevention programme, with school mental health support staff being most commonly selected (80.52%), followed by art teachers (67.53%). More respondents (61.04%) indicated external providers should support such a prevention programme, but higher proportions selected school-based staff roles for programme support. External providers were more frequently recommended to deliver and support an art and wellbeing programme by external staff themselves (Table 2). These findings regarding arts-based programmes were in the context of frequent conversations with students about their mental health, occurring at least once a week for 77.92% of survey participants (73.69% internal staff, 90% of external staff). The majority of respondents (97.40%) felt at least somewhat confident talking to students about mental health problems. However, the amount of training to support student mental health varied across the sample and 59.74% respondents noted additional mental health training needs. The majority of survey respondents (88.16%) thought an art programme should be delivered to smaller groups rather than whole classes. Responses on when the programme would be best delivered were mixed. Less than half survey respondents (41.56%) indicated after school, with around half (50.65%) suggesting in timetabled ‘Personal, Social, Health and Economic’ or health lessons, or as a half day activity (48.05%).

### Qualitative survey and interview results

We constructed three themes from the qualitative data: (1) art is a sanctuary in the school day, (2) resources in schools are stretched, and (3) integration, flexibility and relationships as factors for success. Themes and subthemes are presented in Figure 1, with example quotes in S4 Table.

**Figure 1.**
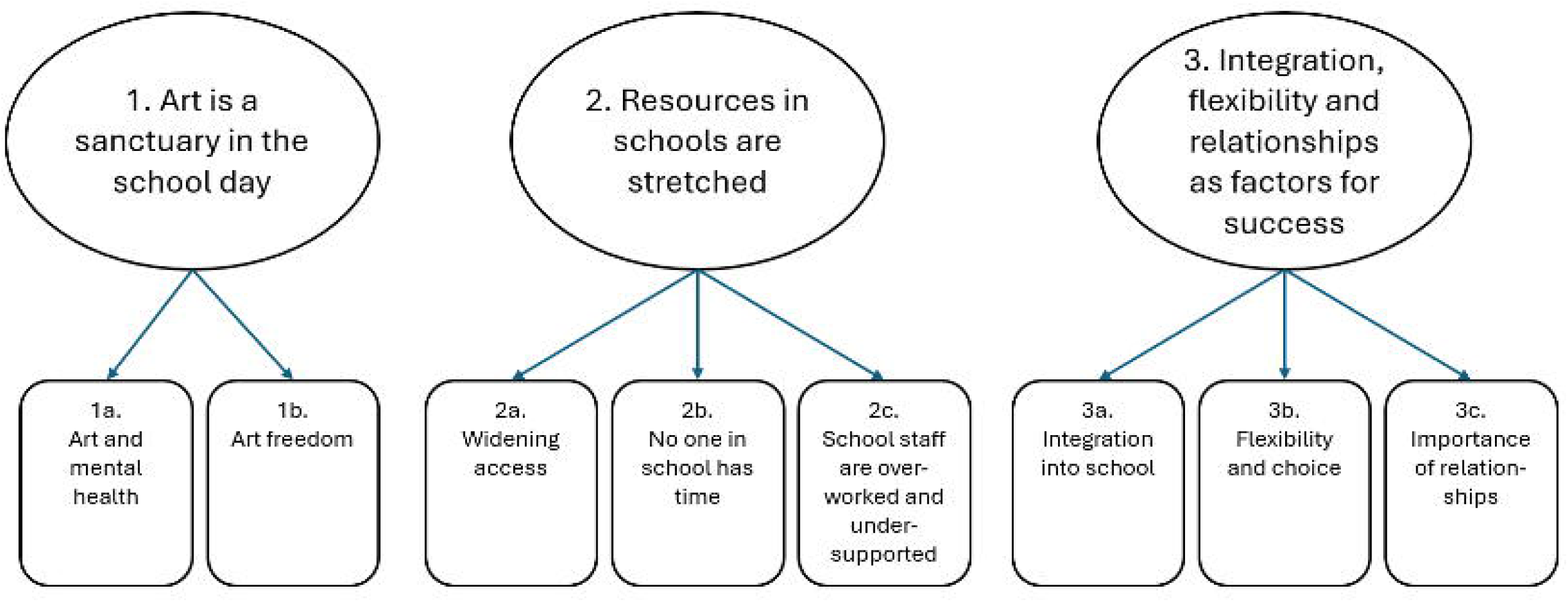
Themes and subthemes constructed from qualitative data.

### Art is a sanctuary in the school day

The first theme considers the role of art in the school setting, with participants emphasising that art provides more than a lesson for young people, even being seen as a sanctuary in the school day. We split this theme into two subthemes: ‘Art and Mental Health’ and ‘Art Freedom’.

#### Art and mental health

Staff discussed that art is intrinsically and uniquely linked to mental health: *“I think art itself lends itself to that [mental health], doesn’t it? That it’s kind of a therapeutic lesson”* (Interview, Art-Teacher-11). Art was seen as different to other subjects because it teaches more than technical or academic skills: *“What is it that art gives to a child, that nothing else does on the curriculum? And it is that place to explore, and experiment safely, and not worry about the outcome”* (Interview, Head-of-Art-1). Furthermore, being creative was seen as a way to help students relax, give a calming space and an opportunity to regulate: *“our inclusion room is a space that a lot of students with special educational needs use, but we also use it as a quiet space for our students with anxiety, or our students who get overwhelmed quite a lot within the school day […] and a lot of the time I walk in and they’re drawing, or they’re painting, or they’re making little models out of things, and it’s all quite creative to keep their minds busy”* (Interview, Pastoral-Team-9). Many participants felt that art facilitated communication by providing a distraction or opening to discuss challenging topics:

*“The conversations around mental health can be really, you know, make them feel really vulnerable. So having something to sort of, distract whilst having those conversations really open - opens them up. […] I use [art and creative methods] all the time in the way of just making the atmosphere a little bit more therapeutic.”* (Interview, EMHP-3).

*“The most genuine kind of conversations I’ve had with [students] is by sitting by their side and I will draw, I will do the same task as they do.”* (Interview, Head-of-Art-2).

The inherent link between art and mental health led to ad hoc clubs being organised and facilitated by invested staff: *“I have been running art well-being sessions in my school two days a week where small groups of learners can create art while discussing problems and socialising with new people.”* (Survey, Art-Teacher-10). Combining art and mental health through a novel programme was therefore seen positively due to the perceived strong link, but programmes should be clearly distinguished from standard art lessons: “*Sometimes non-specialists talk about not being good at art and can ruin the work of art teachers…staff need to be aware of the difference between art (building subject knowledge and skills) and art for mental health development”* (Survey, Art-Teacher-2).

#### Art freedom

Participants saw potential benefits for students engaging in the artistic process freely and with choice. However, ‘art freedom’ can be constrained in school environments because of marking and curriculum pressures and the pressure to do ‘good art’:

*“I think that making art promotes positive mental health when students are able to make art with more flexibility. It is a great way for them to express and communicate without having to use words. However, due to the constraints of the curriculum/number of students in lesson/practicality of getting out different types of equipment etc, it isn’t possible to do that in art lessons”* (Survey, Art-Teacher-9).

Participants discussed how social comparison and the importance placed on the end product can impact students’ ability to enjoy and benefit from the process, with older students particularly affected. Consequently, it would be important for any art and wellbeing programme to communicate with young people that the process is important, not the outcome: “*Because when they go to art class in a school, you know, they’re told what to do, you know, and they compare each other, they compare their work to other’s work, you know, so there’s that separation, you know, trying to separate school art from wellbeing art”* (Interview, Counsellor-3).

*“You don’t have to be a great footballer to enjoy football, you don’t have to be a great artist to enjoy drawing something, you know? You can just have a go and enjoy it”* (Interview, Pastoral-Team-4).

This was noted as challenging in school environments where art freedom is not the norm: *“I have to kind of make clear that I, I’m not there in an academic sense. I’m not there to teach them how to do something, but actually this is a space for them to use, in whichever way they want and I suppose they’re not used to having so much freedom [laughs]”* (Interview, Creative-Arts-Therapist-2).

### Resources in schools are stretched

The second theme encompasses discussions of the pressure in school environments and the restriction of available resources in three subthemes: ‘Widening support’, ‘No one in school has time’, and ‘School staff are overworked and under-supported’.

#### Widening support

Many participants highlighted that student mental health and school attendance had worsened following the COVID-19 pandemic, resulting in reduced resourcing, limited to those with the highest need: *“Because it’s so busy – we’re so busy firefighting, not noticing the little fires that have started, and then turned into a, you know, massive fire. We’re just – we’re just leaving it all until it’s too late”* (Interview, Other-4). Participants discussed the need to widen the support within schools: *“I think it’s important for everybody to have access to do it, and be able to take part in it, because you never know when they might be affected by something that does affect their mental health”* (Interview, Head-of-Art-8) and viewed creating initiatives such as an art and wellbeing prevention programme as a positive step.

Nevertheless, there was a tension between prevention for all and providing a safe space that encouraged engagement within sessions: *“I think students would feel more comfortable to express themselves in smaller groups. However I think it’s important to equip all students with the tools to deal with any mental health issues they may develop”* (Survey, Head-of-Art-10).

#### No one in school has time

Participants highlighted significant time pressures affecting staff and students, with every minute of the day allocated: *“A school day is tightly governed by the clock, tightly governed by, ‘You’ve gotta be here, you’ve gotta be there,’ and you don’t – you don’t get kids in school with free time, and you don’t get adults with free time.”* (Interview, SENCO-4). This environment would lead to practical barriers of taking students and staff off timetable for a prevention programme: *“A counsellor would make sure that their client is ‘safe’ to leave the session but educators would be unable to do this due to time constraints and a lack of training”* (Survey, Art-Teacher-9).

*“I feel that art teachers would be fantastic at helping with supporting this but this would need to be factored into timetabling as most teachers have a very heavy workload”* (Survey, Art-Teacher-9).

Participants noted the need to work with each school on a case-by-case basis to consider what staff would be taken away from to support and deliver the programme and if they would have time: “*All staff in schools are allocated roles, asking staff to step out of role to do something different causes disruption and knocks onto other staff”* (Survey, SENCO-4).

#### School staff are overworked and under-supported

Increased expectations of staff was a recurring concern, with many teaching staff reporting a blurring of roles across education and student mental health support: *“Rather than a role that is guiding them to create the best possible image, you’re stepping into a role that supports them in how they’re feeling, and accepting that the way they want to make imagery is the best way for them to journey through that. … The number of conversations that I’ve had with students over the years about different issues that could or do affect their mental health is too many to count”* (Interview, Head-of-Art-8).

While some participants felt mental health support came as part of their role, some participants identified a need for training to support student’s mental health: *“We haven’t actually looked specifically at mental health this year, even though we’ve got more students school refusing and off for mental health reasons than ever before, so it feels a bit odd that we’re not getting more training, to be honest”* (Interview, Head-of-Art-8).

Improving the level of support for facilitating staff was seen as key for implementing an art and wellbeing programme, with many participants indicating a need for clinical supervision similar to that received by mental health professionals: *“I think It’s important to recognise that art therapy and similar programs can be emotionally demanding, and those involved need to be prepared for this aspect of the work, so the appropriate training and supervision would be needed”* (Survey, Other-9). Support in the form of protected staff time and an adequately resourced programme would reduce the burden of programme delivery and support its success and sustainability: *“I think that as teaching staff we are so busy with day to day school life but having a external support for planning to deliver a programme and prepared resources and materials would be great”* (Survey, Art-Teacher-2).

### Integration, flexibility and relationships as factors for success

The final theme considers factors that are essential to overcome the challenges that may be faced by a potential new art and wellbeing programme in three subthemes: ‘Integration into Schools’, ‘Flexibility and Choice’ and ‘Importance of Relationships’.

#### Integration into school

Participants discussed that previous mental health initiatives they experienced had been short-lived and failed to integrate with the school’s practices. Introducing a programme in line with the schools’ established safeguarding policies, slowly, with good communication and training was seen as a route to promote integration: *“Where we’ve had success with a lot of the changes that we’ve made over the last five, if not ten years, is it’s almost been like a slow build”* (Interview, Other-13). Importantly, to improve integration, participants felt an art and wellbeing programme should not be tokenistic: *“I don’t ever wanna do something in my lesson that’s just ticking a box. If I’m going to do it I want to do it right and really embed it fully”* (Interview, Head-of-Art-2).

Simultaneously, to ensure buy-in from senior leadership, it was seen as important that a programme can meet targets and goals, or be viewed positively by government bodies, such as the UK Office for Standards in Education, Children’s Services and Skills (Ofsted): *“If you get a positive evaluation for a programme, the school, we’re going to carry it on because a, it sounds really cynical now, it looks really good for Ofsted, because they’ve done this programme and it’s been successful, and what have you, if they didn’t carry it on Ofsted, they’d be saying, ‘Well, why have you not carried it on? If it was so successful?’ So they kind of have to carry it on”* (Interview, Mental-Health-Lead-2).

Participants felt that staff ownership of a programme would support its integration and sustainability. They proposed having multiple staff responsible for organising and promoting the programme internally according to the schools’ culture, ensuring the programme continues should a member of staff leave: *“I can highlight two or three people that … would ensure that that happened. And then almost that long term, can they train the next group of people? … And if we talked about that observing and kind of relearning from each other, that, that cycle just continues? So I think on the ground is where sustainability happens in schools”* (Interview, Mental-Health-Lead-3).

#### Flexibility and choice

The views of participants varied widely in how an art and wellbeing programme would look due to schools being diverse, each with their own schedules, practices and cultures. A programme would therefore need to provide flexibility and choice for schools to be successful as a ‘one size fits all’ approach would not be appropriate: *“Each school’s quite different. So some schools want you to have everything done the way that it’s done in their school and some schools are more open to it being done the way it’s done in your service”* (Interview, Creative-Arts-Therapist-1).

Giving staff and students the ability to make an informed choice to take part, depending on their workload and interests, was seen as key for a programme’s success: *“You know, teachers are different [laughs] and some people are so set in their ways, but some people would love it”* (Interview, Art-Teacher-5).

“*Because, a lot of the time, I think, with educational stuff, like if they’re doing, like, lunchtime groups, or after school clubs, it’s kind of forced upon them by teachers or parents, so just being like, ‘This is up to you, if you wanna come,’ kind of shows that, you know, they have that – that kind of control, and choice over it”* (Interview, EMHP-6).

#### Importance of relationships

This sub-theme acknowledges the benefits and challenges of existing relationships, and the importance of building new relationships between staff, external providers and students.

Existing relationships between programme facilitators and students were seen as beneficial to provide programme legitimacy, a contact outside of sessions, and a familiar face: *“I think you would definitely want someone in the room that they were familiar with. Someone who is a reassuring face, a head of year or an art teacher that they’re familiar with or even just seeing the adults in the room”* (Interview, Head-of-Art-2). However existing relationships could also perpetuate pre-existing barriers between teachers and young people, or blur the boundaries between the normal lessons and the programme, thereby impacting its delivery: *“I would say that if an art teacher is in that position, they need to make it clear on their, their role in that group is not kind of, like, creating good art, or making… I suppose establishing boundaries between what’s art class and what this group… Just for clarity for the young people”* (Interview, Creative-Arts-Therapist-2).

Participants perceived involving external professionals as an effective method to create separation from the school day and be an opportunity to create new therapeutic relationships: *“I think it’s good for me, as well, like, external, because I can come in, they don’t know me, and I can build those really, like, positive relationships from the start”* (Interview, EMHP-6).

## Discussion

This mixed-methods study explored experiences of staff working in UK secondary schools around using visual art to promote student mental health and aimed to understand what would be needed to facilitate a school-based visual arts programme to prevent anxiety and depression. Participants felt positively towards using visual arts to support mental health and prevent mental health difficulties in schools and frequently used the arts to support student mental health in practice. Visual art was viewed as strongly and uniquely linked to mental health, and participants felt that enabling art freedom within a school context can be beneficial for mental health when curriculum and marking constraints are lifted. Nevertheless, challenges of implementation were noted, with school staff and resources being stretched, and increases in demand for student mental health support requiring school staff to provide support with limited support and training. Participant views were mixed on how best to deliver an art and wellbeing programme in terms of when it should be delivered and who should deliver and support the programme, highlighting a “one size fits all” approach may not be appropriate. Our study indicates several ways to overcome these challenges including ensuring programme integration into schools, enabling flexibility and choice, and building effective relationships. These factors should form the groundwork for the development of future programmes using arts to support mental health in schools.

In both the quantitative and qualitative data, art was perceived as important for mental health in schools. Almost a third of participants reported using the visual arts at least weekly to support student mental health and promote mental health conversations. These attitudes and practices align with the growing evidence base indicating engagement in the arts can be beneficial for young people’s mental health and wellbeing (14–17) and echoes recent research highlighting that young people aged 16-25 believe art to be essential for mental health (49). In our study, ‘art freedom’, or engaging in the creative process freely and with choice, was perceived as particularly beneficial. This focus on process and journey, rather than output, is often central to creative arts therapies but challenging in a school context with social comparison and a focus on achievement. Our study highlights the importance of including art freedom and choice in future programme designs, and the need to disentangle art for wellbeing from art as an academic subject. We found the majority of work combining the visual arts and mental health occurred informally in schools, rather than as stand-alone programmes. Given school staff welcomed the idea of a preventative programme for mental illness within schools, such a programme could help fill the gap between the perceived importance of visual arts and their structured use. With staff already combining art and mental health in schools, and being positive about a potential formalised programme, school staff could act as effective champions for arts-based programmes.

While there was endorsement of a universal preventative approach to poor student mental health, participants also acknowledged practical constraints and pressures that only allow support for students with the highest mental health needs. The lack of time, resources, and programme integration into schools described by our participants are well-documented barriers for mental health interventions in schools (39,40). Most of our participants felt confident having conversations with students on mental health, but almost 60% reported additional training needs and interview participants noted they would need training to successfully integrate a programme into school practice. School mental health interventions with sufficient training for those delivering them are often more effective and more sustainable (39), and the World Health Organization also stresses the importance of good training (8). Providing adequate training and support for staff delivering programmes is challenging in time-pressured school environments. Participants in the current study suggested that external support and ready-made, well-prepared resources would reduce potential burden for staff delivering programmes. While the evidence on the effectiveness of arts-based prevention programmes for mental health is in its infancy, other prevention and early intervention strategies have been shown to improve young people’s mental wellbeing and reduce costs associated with later mental health difficulties (50,51). Further research into the efficacy and cost-effectiveness of arts prevention programmes is therefore warranted.

Echoing young people’s recommendations to embed arts programming in schools (49), our study found school staff saw integration in schools as critical for the success of arts programmes. Participants highlighted the need for a slow build, good communication and ownership of the programme to promote integration with schools. Good communication with the whole school community can improve buy-in and engagement with prevention programmes (52). Participants in the current study indicated that different strategies may be required to ensure integration and support from staff at different levels in the school system: leadership would benefit from evidence of a programme meeting targets and aligning with regulatory frameworks, whereas staff delivering the programme need their involvement to be meaningful and not seen as a tick-box exercise. School and staff ownership was seen as a key facilitator for programme integration, echoing previous research that found staff excitement and engagement can aid successful implementation of school mental health programmes (53). Based on our findings, there is a need to balance provision of a well-prepared programme that reduces staff burden with flexibility and choice to facilitate ownership.

Our quantitative results highlighted mixed views around who should deliver a programme and when it should be delivered. Exploring these perspectives in interviews highlighted the need for programme flexibility and choice. This may be achieved by allowing self-selection for students and facilitators and enabling flexible implementation, while retaining core elements of the programme and programme fidelity. Similar to previous findings that school pastoral and support staff can encourage student engagement in mental health interventions due to perceived trustworthiness (54), our participants recognised internal staff can provide legitimacy, reassurance and an ongoing contact point for students. Participants also acknowledged the benefits of building new relationships with external staff. Our results suggest that allowing each school and individuals to choose who is best placed to support and deliver an art and wellbeing programme will likely increase its chances of success, but that a combination of school staff and external providers can be beneficial. The addition of external staff could also provide additional support for school staff facilitators, or empower them to be able to run the programme.

Despite the desire for a universal preventative approach evident in the qualitative data, the quantitative data indicated a preference for small group delivery over whole class delivery. Maintaining smaller groups limits the ability to apply a whole school approach and can increase programme resourcing costs. Reasons for choosing to restrict group size centred around student comfort and viewing a smaller group as more conducive for wellbeing discussions. This may be one reason prevention approaches seem promising in small group contexts but fail to replicate in large-scale trials (11). Prevention in schools may therefore benefit from a multi-level approach, that enables universal access for the whole school, smaller groups that feel safer for students, and work at the individual level for those with higher needs. For an arts prevention programme, this may look like regular arts activities accessible to a whole year group through half-day events or tutor time, smaller weekly group arts workshops after school or off-timetable for students who want to engage more and are at risk of developing mental health issues, and one-to-one sessions with a trained creative therapist for individuals with greater mental health needs. School prevention programmes that have trialled multi-level (non-arts) approaches have indicated reduction in depressive symptoms (for example, the SEHER trial (55)), but need extensive integration and maintained funding to be sustainable (56).

Based on our findings, we provide recommendations for teams developing arts-based mental health interventions and prevention programmes in schools in Table 3.

**Table 3.** Recommendations for developing school art-based mental health interventions.

| **Number** | **Recommendation** |
|---|---|
| 1 | Build flexibility within the intervention for students to choose activities and allow them to be creative in their own way. It is this creative freedom, without pressure, that facilitates wellbeing conversations and may confer beneficial effects. |
| 2 | Intervention implementation will need to be flexible so it can be tailored to the needs and contexts of each school. |
| 3 | Mental health intervention delivery can benefit from internal school staff who know students and can maintain links, and external professionals who bring additional expertise and can create new, positive relationships with students. |
| 4 | Appropriate training and support for those delivering the intervention is essential. If school staff are delivering the intervention, then training and support will likely need to be in addition to what is already available; for instance, clinical supervision, similar to what mental health professionals receive. |
| 5 | Reduce the burden of intervention delivery with ready-made resources and session plans. |
| 6 | Programme integration in schools can be promoted through school ownership and staff champions, clear communication, adequately-timed lead in before the intervention starts, sustained long-term delivery, and alignment with regulatory frameworks (e.g. Ofsted). |
| 7 | Participation in intervention delivery and support (staff) or as a beneficiary (student) should be a choice made by each individual. |

### Strengths and limitations

We used a mixed-methods design, collecting and analysing rich qualitative data and broader quantitative data. This complementary data analysis enabled a breadth of views to be collected but also in-depth insights in this complex research field. However, the time and resource needed to conduct interviews meant that less time was spent on recruitment and may have resulted in a smaller quantitative sample than if the study had employed surveys alone. While we incorporated the views of a diverse range of professions working within schools in the survey, the smaller sample size and number of job roles precluded the use of inferential statistics. Moreover, most of the sample worked in English schools, therefore the findings may not be relevant to different educational contexts. The survey and interview samples also comprised staff who would likely be involved in the support or delivery of an art and wellbeing programme. These staff are more likely to have positive attitudes towards arts and mental health initiatives than broader staff groups (for instance, teachers specialising in non-arts subjects) and school communities. Therefore, further research is required to ascertain acceptability of arts-based programmes more broadly with school staff with different roles (including school leaders), as well as parents and students. Given the potential importance of art freedom in promoting student mental health, additional research to elucidate facilitators to art freedom in a school context is also warranted to improve effectiveness of school-based arts programmes.

### Conclusions

This mixed-methods study found that visual arts were seen as important for supporting and promoting student mental health and were used frequently used for this purpose, despite most staff receiving no training for these activities. A visual arts programme to help prevent anxiety and depression within schools was seen as a positive initiative but would encounter implementation challenges. Our findings indicate that a number of factors are needed to successfully integrate an art and wellbeing programme into a busy school context including art freedom, flexibility, choice, a combination of internal and external programme facilitators, appropriate training and supervision for facilitators, ready-made resources, and school ownership. Future development of visual art prevention programmes should take these factors into account to ensure successful and sustainable implementation.

## Data Availability

Supporting data has been published under access restrictions due to ethical considerations. Details on how to access data are available at the University of Bristol data repository, data.bris, under the DOI https://doi.org/10.5523/bris.3alr183jg9nbs2w5ab8xhxfyfg.

## Acknowledgements

We gratefully acknowledge the participants for sharing their experiences and their time with us for this research.

## Supporting information

**S1 Table.** Good Reporting of A Mixed Methods Study (GRAMMS) checklist

**S1 Figure.** Participant recruitment flow chart

**S2 Table.** Individual participant backgrounds

**S1 File.** Full survey text

**S2 File.** Interview topic guide

**S3 Table.** Reflexive position of team

**S4 Table**. Themes and example quotes

