## Supplementary material for "Visual art for the prevention of student mental health difficulties in schools: Mixed-methods analysis of staff surveys and interviews": S1 File

### S1 File. Full survey text

Below are the questions asked within the online survey. In response to these questions, either multiple options were given or a space was provided for an ‘open text response’. Where the option ‘other’ was provided, a subsequent open text response box was also provided. Questions with options provided were either single forced-choice questions or multiple-choice questions (where multiple answers could be selected). Multiple choice questions are indicated by (MCQ). An asterisk (*) denotes items not included in analysis or available data.

#### About you

**How would you describe your gender?**

- Male
- Female
- Non-binary
- Other (please specify)

**How old are you?**

- 18 to 24
- 25 to 29
- 30 to 34
- 35 to 39
- 40 to 44
- 45 to 49
- 50 to 59
- 60 to 64
- 65+

**How would you describe your ethnicity?**

- Asian or Asian British: Indian
- Asian or Asian British: Pakistani
- Asian or Asian British: Bangladeshi
- Asian or Asian British: Chinese
- Asian or Asian British: Any other Asian background
- Black, Black British, Caribbean or African: Caribbean
- Black, Black British, Caribbean or African: African
- Black, Black British, Caribbean or African: Any other Black, Black British, or Caribbean background
- Mixed or multiple ethnic groups: White and Black Caribbean
- Mixed or multiple ethnic groups: White and Black African
- Mixed or multiple ethnic groups: White and Asian
- Mixed or multiple ethnic groups: Any other Mixed or multiple ethnic background
- White: English, Welsh, Scottish, Northern Irish or British
- White: Irish
- White: Gypsy or Irish Traveller
- White: Roma
- White: Any other White background
- Other ethnic group: Arab
- Other ethnic group: Any other ethnic group

**How did you hear about this survey?***

Open text response

#### About your professional background

**What region do you work in?** (MCQ)

- England
- Wales
- Scotland
- Northern Ireland

***What is the name of your school? We ask this so we can count how many different schools are represented in the research. We will not publish or share the school name.***

*Open text response*

***Is this a fee-paying school?***

- No
- Yes

***Does the school you work in fall under an academy or trust?***

- No
- Yes

***What is the name of the academy/trust your school is a part of? We ask this to understand the school context. We will not share or publish this information.***

Open text response

**How would you describe your professional background?**

- Art teacher
- Head of Art or Curriculum Leader for Art
- Leadership role for mental health (e.g. Designated Senior Lead for Mental Health, Lead Practitioner for Health and Wellbeing)
- Member of the pastoral team
- Teaching assistant
- School counsellor
- Creative arts therapist
- Thrive practitioner
- Emotional Literacy Support Assistant (ELSA)
- SENCO (Special Educational Needs Coordinator) or ALNCO (Additional Learning Needs Coordinator)
- Educational Mental Health Practitioner
- Other (please specify)

**How long have you worked in an educational setting?**

- Less than a year
- 1-4 years
- 5-9 years
- 10+ years

**How much do you currently use the arts in your work? (e.g. creative writing, experiencing music, creating images)**

- None
- A little
- Some
- A lot

**How much do you currently use visual arts in your work? (e.g. creating images or sculptures, drawing, painting)**

- None
- A little
- Some
- A lot

#### Pupil mental health

**Do you feel that, in your role, you should be helping pupils with their mental health problems?**

*Open text response*

**In the past year, how often have you had conversations with pupils about mental health and wellbeing?**

- Never
- Less than once a month
- Once a month
- 2-3 times a month
- Once a week
- 2-4 times a week
- Once a day
- Multiple times a day

**How confident do you feel talking to pupils about mental health problems?**

- Not at all confident
- Somewhat confident
- Moderately confident
- Extremely confident

#### Mental health training

**Over the past 3 years, how many days of training have you had to support pupils with their mental health?**

- None
- Less than a day
- 1 day
- 2 days
- 3 days
- 4 days
- 5 days
- 6+ days

**What was that training?**

*Open text response*

**Are there any mental health training needs that you have identified that you would like support with?**

- No
- Yes

**If yes, please describe the training needs that you would like to be met**

*Open text response*

**Have you had any training on supporting pupil mental health that has incorporated creativity or use of the arts? (e.g. creative writing, experiencing music, creating images)**

- No
- Yes

**If yes, please describe the training.**

*Open text response*

**Have you had any training on supporting teacher/staff mental health that has incorporated creativity or use of the arts? (e.g. creative writing, experiencing music, creating images)**

- No
- Yes

**If yes, please describe the training.**

*Open text response*

#### Art and mental health

**In your opinion, how important is creating visual art (e.g. creating images or sculptures, drawing, painting) for supporting pupil mental health?**

- Very unimportant
- Unimportant
- Neutral
- Important
- Very important

**What are your reasons for this choice?**

*Open text response*

**In the past year, how often have you used visual art as a tool to promote or support pupil mental health in your school? (e.g. working with pupils to create art to promote mental health awareness, using art therapeutically to support pupils with mental health problems)**

- Never
- Less than once a month
- Once a month
- 2-3 times a month
- Once a week
- 2-4 times a week
- Once a day
- Multiple times a day

**What types of art activities have you used to promote or support pupil mental health?**

*Open text response*

#### Developing an art-based programme in schools

**What resources (such as materials, training, or support) do you think would be helpful in incorporating visual art into a programme to help prevent anxiety and depression in pupils?**

*Open text response*

**What staff should be involved in supporting an art-based programme to help prevent pupil anxiety and depression? (please tick all options that you think apply)** (MCQ)

- School mental health support staff
- Leaders for mental health (e.g. Designated Senior Lead for Mental Health, Lead Practitioner for Health and Wellbeing)
- Heads of Art or Curriculum Leaders for Art
- The pastoral team
- Art teachers
- Non-art teachers
- Teaching support staff
- External providers/specialists
- Other (please specify)

**If you would like to expand upon the reasons for your decision, please do so here:**

*Open text response*

**What staff should be involved in delivering an art-based programme to help prevent pupil anxiety and depression? (please tick all options that you think apply)** (MCQ)

- School mental health support staff
- Leaders for mental health (e.g. Designated Senior Lead for Mental Health, Lead Practitioner for Health and Wellbeing)
- Heads of Art or Curriculum Leaders for Art
- The pastoral team
- Art teachers
- Non-art teachers
- Teaching support staff
- External providers/specialists
- Other (please specify)

**If you would like to expand upon the reasons for your decision, please do so here**

*Open text response*

**Do you think an art programme should be aimed at whole classes or smaller groups? (For instance, smaller groups of pupils at high risk of developing anxiety/depression, or pupils who want to join the group)**

- Whole classes
- Smaller groups

**If you would like to expand upon the reasons for your decision, please do so here**

*Open text response*

**At what time during the school day would an art programme be best delivered?**

- PSHE or health lessons
- Art lessons
- Tutor time
- Half day activity
- Whole day activity (e.g. off-timetable day, enrichment day, or drop down day)
- Extra curricular club after school
- Extra-curricular club during lunch time
- Other (please specify)

#### Safety and boundaries

**Do you have any concerns about a programme using art to help prevent anxiety and depression?**

*Open text response*

**Helping pupils to manage their emotions and find creative ways to express themselves may bring up unpleasant thoughts and feelings for some people. Do you think that school staff will be able to cope with feelings that arise within them and continue to support pupils for the duration of the session?**

*Open text response*

**Some experiences within the programme could leave a lasting impression. Do you think that school staff will be able to handle their own feelings following a session? (e.g. will school staff be able to "switch off" when they get home?)**

*Open text response*

#### Any other comments

**Are there any other comments or thoughts you would like to share? You can use this space to share anything you feel we should have asked about or let us know any reasoning for previous answers.**

*Open text response*
