## Supplementary material for "Visual art for the prevention of student mental health difficulties in schools: Mixed-methods analysis of staff surveys and interviews": S1 Table

### S1 Table. Good Reporting of A Mixed Methods Study (GRAMMS) checklist

O’Cathain A, Murphy E, Nicholl J. The quality of mixed methods studies in health services research. J Heal Serv Res Policy. 2008;13(2):92–8.

| **Guideline** | **Section & Page** |
| --- | --- |
| Describe the justification for using a mixed methods approach to the research question | Introduction: The current study  Pages 4-5 |
| Describe the design in terms of the purpose, priority and sequence of methods | Methods: Study Design  Page 5 |
| Describe each method in terms of sampling, data collection and analysis | Methods: Participants and recruitment; Data collection; Data analysis  Pages 5-13 |
| Describe where integration has occurred, how it has occurred and who has participated in it | Methods: Data analysis  Pages 12-13 |
| Describe any limitation of one method associated with the present of the other method | Discussion: Strengths and limitation  Page 32 |
| Describe any insights gained from mixing or integrating methods | Discussion  Pages 27-32 |
