## Supplementary material for "Visual art for the prevention of student mental health difficulties in schools: Mixed-methods analysis of staff surveys and interviews": S2 File

### S2 File. Interview topic guide

| **Introductory questions** | |
| --- | --- |
| What comes to mind when you think of visual art in the context of mental health or wellbeing? OR  What are your thoughts on the use of visual art in the context of mental health or wellbeing? | |
|  | *PROMPT:* Do you have experience of art being used in a mental health or wellbeing context, for example in your work in educational settings? |
| Do you have any initial thoughts on the idea of a school-based programme (series of lessons) around art and mental health which aims to prevent problems such as depression and anxiety in young people? | |
| What do you think would an art and wellbeing programme like this would look like? E.g. what materials being used, how many pupils in the class, how many facilitators, how long are the sessions, | |

| **Who should be involved in the art-based programme?** | | |
| --- | --- | --- |
| Who do you think should be involved in leading and supporting an art programme for preventing anxiety and depression in schools? | | |
| What role do you think *art teachers/school staff who support mental health** would have in supporting programme around art and mental health? | | |
|  | *PROMPT/FOLLOW UP:* Would *art teachers/school staff who support mental health* have a role in leading these sessions? | |
|  | *PROMPT/FOLLOW UP:* What support would be needed for *art teachers/school staff who support mental health* to lead these sessions? Within the sessions themselves and support outside of the sessions | |
|  | *PROMPT/FOLLOW UP:* What training on mental health do *art teachers* currently have? What training would be needed? | *PROMPT/FOLLOW UP:* What training on art and creativity do *school staff who support mental health* currently have? What training would be needed? |
| What are your views on art teachers’ roles in pastoral care? | | |
|  | *PROMPT/FOLLOW UP:* Is supporting an art-based programme something that art teachers would want built into their role? | |
| Can art teachers develop spaces for pupil self-expression that don’t focus on academic attainment? | | |
| Would *art teachers/school staff who support mental health* have time/capacity to spend on training and delivering an art programme? | | |

**Sustainability**

What would be needed to ensure an art prevention programme can be useful beyond the lifespan of the research study?

What support would schools need to run the programme over longer term? E.g. place to offload, supervision/sign-posting protocol, teacher support network, ongoing training

| **Implementation** | |
| --- | --- |
| What time of school day would the programme be best delivered? e.g. PSHE lessons, art lessons, after school, lunch break, tutor time, off-timetable days or half-days | |
|  | How long should the programme be delivered for? |
|  | What support should be provided for pupils on completion? E.g. sign-posting, additional sessions |
| Should the prevention programme cover whole classes or should we consider smaller groups? | |
|  | *IF SMALLER GROUPS:* How should we decide who goes in these smaller groups? (e.g. self-selection, high-risk pupils) |
| How can we encourage students to engage in the process of the art programme? | |
|  | *PROMPT:* What can be done in sessions to make the art programme as inclusive as possible? |
| How can we create a safe space in the classroom where shared communication and trust can flourish? | |
|  | *PROMPT:* What are the possible barriers to pupil’s feeling safe in the sessions? |
| What materials do you think would be used in the art programme? | |
|  | PROMPT: Where would you get these materials? E.g. school already supplied, extra resource |

| **Concerns and safety** | |
| --- | --- |
| Are there any concerns you might have about a programme using art to prevent pupil anxiety and depression? | |
| Do you think school staff will be able to safely deliver a programme to help pupils manage their emotions and find creative ways to express themselves? E.g. boundaries with other pupil’s/confidentiality, attending to needs of all pupils in the class, pupils feeling vulnerable/exposed, not doing psychological harm | |
| How will school staff be able to cope with feelings that arise within them and continue to support pupils for the duration of the session? E.g. dealing with triggers, handle emotions | |
| How will school staff be able to handle their own feelings following a session? (e.g. will school staff be able to "switch off" when they get home?) | |
| Does your school have a policy covering student mental health? | |
|  | *PROMPT/FOLLOW UP:* What does the policy cover? How would an art programme to prevent anxiety and depression fit with this? |

**Final points**

Is there anything you thought I would ask you but haven’t?

Is there anything else you would like to share around what we have been talking about?

Do you have any questions?
