## Supplementary material for "Visual art for the prevention of student mental health difficulties in schools: Mixed-methods analysis of staff surveys and interviews": S2 Table

### S2 Table. Individual participant backgrounds

| ***Pseudonym*** | **Interview** | **Background** | **Gender** | **Ethnicity** | **Age** | **Region of work** |
| --- | --- | --- | --- | --- | --- | --- |
| *Art-Teacher-1* | Y | Art teacher | Female | White: Any other White background | 30 to 34 | England |
| *Art-Teacher-2* |  | Art Teacher | Female | White: English, Welsh, Scottish, Northern Irish or British | 40 to 44 | England |
| *Art-Teacher-3* |  | Art Teacher | Female | White: English, Welsh, Scottish, Northern Irish or British | 25 to 29 | England |
| *Art-Teacher-4* |  | Art teacher | Female | White: Irish | 25 to 29 | England |
| *Art-Teacher-5* | Y | Art teacher | Female | White: Irish | 45 to 49 | England |
| *Art-Teacher-6* |  | Art teacher | Female | Mixed or multiple ethnic groups: White and Asian | 35 to 39 | England |
| *Art-Teacher-7* |  | Art teacher | Female | White: English, Welsh, Scottish, Northern Irish or British | 40 to 44 | England |
| *Art-Teacher-8* |  | Art teacher | Female | White: English, Welsh, Scottish, Northern Irish or British | 25 to 29 | England |
| *Art-Teacher-9* |  | Art teacher | Female | White: English, Welsh, Scottish, Northern Irish or British | 30 to 34 | England |
| *Art-Teacher-10* | Y | Art Teacher | Female | White: English, Welsh, Scottish, Northern Irish or British | 45 to 49 | Wales |
| *Art-Teacher-11* | Y | Art Teacher | Female | White: English, Welsh, Scottish, Northern Irish or British | 35 to 39 | England |
| *Counsellor-1* | Y | Counsellor | Female | White: English, Welsh, Scottish, Northern Irish or British | 65+ | Wales |
| *Counsellor-2* |  | School Counsellor | Female | White: English, Welsh, Scottish, Northern Irish or British | 65+ | Wales |
| *Counsellor-3* | Y | School counsellor | Female | White: English, Welsh, Scottish, Northern Irish or British | 50 to 59 | England |
| *Counsellor-4* |  | School Counsellor | Female | White: English, Welsh, Scottish, Northern Irish or British | 50 to 59 | England |
| *Creative-Arts Therapist-1* | Y | Creative Arts Therapist | Female | White: Any other White background | 25 to 29 | Scotland |
| *Creative-Arts Therapist-2* | Y | Creative Arts Therapist | Female | White: English, Welsh, Scottish, Northern Irish or British | 25 to 29 | Scotland |
| *Creative-Arts Therapist-3* |  | Creative Arts Therapist | Female | White: English, Welsh, Scottish, Northern Irish or British | 40 to 44 | England |
| *Creative-Arts Therapist-4* |  | Creative Arts Therapist | Male | White: English, Welsh, Scottish, Northern Irish or British | 45 to 49 | England |
| *ELSA-1* |  | Emotional Literacy Support Assistant | Female | White: English, Welsh, Scottish, Northern Irish or British | 45 to 49 | England |
| *ELSA-2* |  | Emotional Literacy Support Assistant | Female | White: English, Welsh, Scottish, Northern Irish or British | 25 to 29 | England |
| *EMHP-1* | Y | Educational Mental Health Practitioner | Female | Other ethnic group: Any other ethnic group | 45 to 49 | England |
| *EMHP-2* | Y | Educational Mental Health Practitioner | Female | White: English, Welsh, Scottish, Northern Irish or British | 30 to 34 | England |
| *EMHP-3* | Y | Educational Mental Health Practitioner | Female | White: English, Welsh, Scottish, Northern Irish or British | 25 to 29 | England |
| *EMHP-4* |  | Educational Mental Health Practitioner | Female | White: English, Welsh, Scottish, Northern Irish or British | 25 to 29 | England |
| *EMHP-5* |  | Educational Mental Health Practitioner | Female | White: English, Welsh, Scottish, Northern Irish or British | 45 to 49 | England |
| *EMHP-6* | Y | Educational Mental Health Practitioner | female | White: English, Welsh, Scottish, Northern Irish or British | 25 to 29 | England |
| *EMHP-7* | Y | Educational Mental Health Practitioner | female | White: Any other White background | 18 to 24 | England |
| *EMHP-8* | Y | Educational Mental Health Practitioner | female | White: English, Welsh, Scottish, Northern Irish or British | 30 to 34 | England |
| *EMHP-9* |  | Educational Mental Health Practitioner | Female | White: English, Welsh, Scottish, Northern Irish or British | 25 to 29 | England |
| *Head-of-Art-1* | Y | Head of Art or Curriculum Lead for Art | Female | White: English, Welsh, Scottish, Northern Irish or British | 45 to 49 | England |
| *Head-of-Art-2* | Y | Head of Art or Curriculum Lead for Art | Female | White: English, Welsh, Scottish, Northern Irish or British | 30 to 34 | England |
| *Head-of-Art-3* |  | Head of Art or Curriculum Lead for Art | Female | White: Any other White background | 50 to 59 | England |
| *Head-of-Art-4* |  | Head of Art or Curriculum Lead for Art | Female | White: English, Welsh, Scottish, Northern Irish or British | 45 to 49 | England |
| *Head-of-Art-5* |  | Head of Art or Curriculum Lead for Art | Female | White: English, Welsh, Scottish, Northern Irish or British | 30 to 34 | England |
| *Head-of-Art-6* |  | Head of Art or Curriculum Lead for Art | Female | White: English, Welsh, Scottish, Northern Irish or British | 40 to 44 | Wales |
| *Head-of-Art-7* |  | Head of Art or Curriculum Lead for Art | Female | White: English, Welsh, Scottish, Northern Irish or British | 30 to 34 | England |
| *Head-of-Art-8* | Y | Head of Art or Curriculum Lead for Art | Female | White: English, Welsh, Scottish, Northern Irish or British | 40 to 44 | England |
| *Head-of-Art-9* |  | Head of Art or Curriculum Lead for Art | Male | White: English, Welsh, Scottish, Northern Irish or British | 35 to 39 | England |
| *Head-of-Art-10* |  | Head of Art or Curriculum Lead for Art | Female | White: English, Welsh, Scottish, Northern Irish or British | 35 to 39 | England |
| *Head-of-Art-11* |  | Head of Art or Curriculum Lead for Art | Male | White: English, Welsh, Scottish, Northern Irish or British | 40 to 44 | England |
| *Head-of-Art-12* | Y | Head of Art or Curriculum Lead for Art | Female | White: English, Welsh, Scottish, Northern Irish or British | 35 to 39 | England |
| *Head-of-Art-13* |  | Head of Art or Curriculum Lead for Art | Female | White: English, Welsh, Scottish, Northern Irish or British | 35 to 39 | England |
| *Mental-Health-Lead-1* |  | Leadership role for mental health | Female | White: English, Welsh, Scottish, Northern Irish or British | 45 to 49 | England |
| *Mental-Health-Lead-2* | Y | Leadership role for mental health | Female | White: English, Welsh, Scottish, Northern Irish or British | 50 to 59 | England |
| *Mental-Health-Lead-3* | Y | Leadership role for mental health | Female | White: English, Welsh, Scottish, Northern Irish or British | 25 to 29 | England |
| *Other-1* |  | Cover Teacher | Female | White: English, Welsh, Scottish, Northern Irish or British | 35 to 39 | England |
| *Other-2* | Y | Higher level Teaching Assistant for physical and sensory needs | Female | White: Any other White background | 45 to 49 | England |
| *Other-3* |  | Social, Emotional, and Mental Health Practitioner | Female | White: English, Welsh, Scottish, Northern Irish or British | 35 to 39 | England |
| *Other-4* | Y | Specialist Autism Teacher | Female | White: English, Welsh, Scottish, Northern Irish or British | 50 to 59 | England |
| *Other-5* |  | Supported Self-Help Guidance Practitioner | Female | White: English, Welsh, Scottish, Northern Irish or British | 50 to 59 | England, Wales |
| *Other-6* |  | Teaching Assistant, Art Technician, and Creative Arts Therapist | Female | White: English, Welsh, Scottish, Northern Irish or British | 25 to 29 | England |
| *Other-7* |  | Artist, Youth Project Worker | Male | White: English, Welsh, Scottish, Northern Irish or British | 40 to 44 | Wales, England, Scotland, Northern Ireland |
| *Other-8* |  | Inclusion Leader | Female | White: English, Welsh, Scottish, Northern Irish or British | 25 to 29 | England |
| *Other-9* |  | Art Technician | Female | White: English, Welsh, Scottish, Northern Irish or British | 40 to 44 | England |
| *Other-10* |  | Educational Psychologist | Female | White: English, Welsh, Scottish, Northern Irish or British | 30 to 34 | England |
| *Other-11* |  | Senior Educational Psychologist | Female | White: English, Welsh, Scottish, Northern Irish or British | 40 to 44 | England |
| *Other-12* |  | Educational Psychologist | Female | White: English, Welsh, Scottish, Northern Irish or British | 35 to 39 | England |
| *Other-13* | Y | Art Teacher and Head of Year. Former Head of Faculty | Male | White: English, Welsh, Scottish, Northern Irish or British | 40 to 44 | England |
| *Pastoral-Team-1* |  | Member of Pastoral Team | Female | White: English, Welsh, Scottish, Northern Irish or British | 35 to 39 | England |
| *Pastoral-Team-2* |  | Member of Pastoral Team | Female | White: English, Welsh, Scottish, Northern Irish or British | 25 to 29 | England |
| *Pastoral-Team-3* |  | Member of Pastoral Team | Female | White: English, Welsh, Scottish, Northern Irish or British | 40 to 44 | England |
| *Pastoral-Team-4* | Y | Member of Pastoral Team | Female | White: English, Welsh, Scottish, Northern Irish or British | 50 to 59 | England |
| *Pastoral-Team-5* |  | Member of Pastoral Team | Female | White: English, Welsh, Scottish, Northern Irish or British | 40 to 44 | England |
| *Pastoral-Team-6* |  | Member of Pastoral Team | Female | White: English, Welsh, Scottish, Northern Irish or British | 45 to 49 | England |
| *Pastoral-Team-7* |  | Member of Pastoral Team | Female | White: English, Welsh, Scottish, Northern Irish or British | 30 to 34 | England |
| *Pastoral-Team- 8* | Y | Member of Pastoral Team | Female | Mixed or multiple ethnic groups: White and Black African | 50 to 59 | England |
| *Pastoral-Team-9* | Y | Member of Pastoral Team | Female | White: English, Welsh, Scottish, Northern Irish or British | 25 to 29 | England |
| *SENCO-1* |  | SENCO or ALNCO | Female | Mixed or multiple ethnic groups: White and Black Caribbean | 40 to 44 | England |
| *SENCO-2* |  | SENCO or ALNCO | Female | White: English, Welsh, Scottish, Northern Irish or British | 40 to 44 | England |
| *SENCO-3* |  | SENCO or ALNCO | Female | White: English, Welsh, Scottish, Northern Irish or British | 40 to 44 | England |
| *SENCO-4* | Y | SENCO or ALNCO | Male | White: English, Welsh, Scottish, Northern Irish or British | 60 to 64 | England |
| *Teaching-Assistant-1* |  | Teaching Assistant | Female | White: English, Welsh, Scottish, Northern Irish or British | 45 to 49 | England |
| *Teaching-Assistant-2* |  | Teaching Assistant | Female | White: English, Welsh, Scottish, Northern Irish or British | 50 to 59 | England |
| *Teaching-Assistant-3* |  | Teaching Assistant | Female | Asian or Asian British: Indian | 40 to 44 | England |
| *Teaching-Assistant-4* |  | Teaching Assistant | Female | White: English, Welsh, Scottish, Northern Irish or British | 35 to 39 | England |
| *Thrive-1* |  | Thrive Practitioner | Female | White: English, Welsh, Scottish, Northern Irish or British | 30 to 34 | England |

SENCO (Special Educational Needs Coordinator) or ALNCO (Additional Learning Needs Coordinator)
