## Supplementary material for "Visual art for the prevention of student mental health difficulties in schools: Mixed-methods analysis of staff surveys and interviews": S3 Table

### S3 Table. Reflexive position of team

| **Initials** | **Role** | **Reflexivity statement** |
| --- | --- | --- |
| BE | Conducted interviews and thematic analysis | BE is a postdoctoral researcher with a background working in mental health services. She is a white, middle-class woman. Although she does not consider herself particularly artistic, she engages in creative practices such as needlework and textile work, which she views as beneficial to her wellbeing. BE worked on a project led by NW to co-produce an arts and wellbeing programme aimed at preventing anxiety and depression. Because of this work, and her personal experience, she approached the study with the assumption that arts can support wellbeing and mental health. |
| HC | Conducted interviews | HC is a lived experience mental health researcher, alongside her primary work in access to justice. She has supported qualitative analysis across a number of studies, including on novel treatments for anorexia nervosa and psychedelic-assisted therapy for eating disorders and depression. HC is a white, neurotypical cisgender woman in her early thirties. Her personal experience with visual art has been mixed; however, her mother is a visual artist, and HC has a strong appreciation for other art forms, particularly music. As a result, HC approached the research without a settled view of the impact of visual arts on wellbeing and mental health. |
| NW | Project lead, conducted interviews, supervised thematic analysis, conducted quantitative analysis and integration of mixed methods | NW is a mental health researcher with experience primarily in quantitative methods with developing qualitative expertise. Prior to her degrees, NW studied and enjoyed visual art as a subject at school but only engages in visual art sporadically as an adult. She is a white woman of English heritage. She leads a project looking to co-produce and art and wellbeing programme aimed at preventing anxiety and depression. Because of this, NW approached the research with an assumption that visual arts have the potential to be beneficial for wellbeing and mental health. |
| JK | Supervised thematic analysis | JK is a Professor of public mental health. She has led a number of school-based studies focused on mental health improvement and is aware of the challenges schools face in prioritising preventative mental health. JK is a mixed methods but primarily qualitative researcher. She is a middle-aged, middle-class white woman. JK considers herself 'bad' at visual art. However, she is an advocate for arts-based subjects supporting mental health. This is based on her own experience, findings from an earlier qualitative study with secondary school students and the wider literature. |
