## Supplementary material for "Visual art for the prevention of student mental health difficulties in schools: Mixed-methods analysis of staff surveys and interviews": S4 Table

### S4 Table. Themes and example quotes

| **Subtheme** | **Quote** |
| --- | --- |
| **Theme 1: Art is a sanctuary in the school day** | |
| **Art and mental health** | “We do mindfulness activities, creating mindful pictures, giving students the chance to focus on something whilst they are in crisis taking their mind off the problem.” (Survey, Thrive-1) |
|  | “Our inclusion room is a space that a lot of students with special educational needs use, but we also use it as a quiet space for our students with anxiety, or our students who get overwhelmed quite a lot within the school day. And our inclusion manager is not an art teacher, so it’s not any sort of structured art input, but there are resources there that a lot of students do use, and a lot of the time I walk in and they’re drawing, or they’re painting, or they’re making little models out of things, and it’s all quite creative, to keep their minds busy. And it is really widely used by our students who like to have that quiet time” (Interview, Pastoral-Team-9) |
|  | “I think to my students who are going through things or my students that are, you know, behavioural issues, whatever it might be, the most genuine kind of conversations I’ve had with them is by sitting by their side and I will draw, I will do the same task as they do.” (Interview, Head-of-Art-2) |
|  | “I have been running art well-being sessions in my school two days a week where small groups of learners can create art while discussing problems and socialising with new people.” (Survey, Art-Teacher-10) |
|  | “I think art itself lends itself to that [mental health], doesn't it? That it's kind of a therapeutic lesson” (Interview, Art-Teacher-11) |
|  | “I think, as art teachers, we’re in a really privileged position, because we get to know students a lot better than, say, a maths teacher, or a physics teacher, because we are asking them, all the time, to go inwards, and go outwards, and give something of themselves.” (Interview, Head-of-Art-1) |
|  | “Only concern sometimes non-specialists talk about not being good at art and can ruin the work of art teachers…staff need to be aware of the difference between art (building subject knowledge and skills) and art for mental health development.” (Survey, Art-Teacher-2) |
|  | “Art should also not be deskilled into a purely therapeutic- zero skill hobby.” (Survey, Art-Teacher-3) |
|  | “What is it that art gives to a child, that nothing else does on the curriculum?’ And it is that place to explore, and experiment safely, and not worry about the outcome.” (Interview, Head-of-Art-1) |
|  | “Students explore their own ideas through projects, a lot of students choose to explore mental health issues as part of these projects.” (Survey, Head-of-Art-10) |
|  | “I think the resilience it builds enable students to cope better in everyday life. We often find in art that we get to know students more as they will be unable to talk about what they are feeling but will incorporate it into their work. Art lessons are often the only times in a school day where students are able to reflect on their personal lives and connect their interests into school.” (Survey, Art-Teacher-3) |
|  | “The first thing that comes to mind for me, is actually like, building the therapeutic alliance. I think that when I have sessions with kids, especially like, young secondary and primary, actually older secondary as well to be fair, if they’re not doing something like drawing or doodling or fidgeting with something, it’s really hard to build that therapeutic alliance because it’s so intense. The conversations around mental health can be really, you know, make them feel really vulnerable. So having something to sort of, distract whilst having those conversations really open - opens them up. So I think that’s the first thing that, sort of, drew me to doing this research, because you know, I use it all the time in the way of just making the atmosphere a little bit more therapeutic.” (Interview, EMHP-3) |
|  | “I think art teachers, because of the nature of the subject, tend to become more pastorally inclined anyway. Because of the fact art rooms are often open during breaks, and lunch times, and after school, because, in some ways, it can be less academically relaxed than, say, a maths room, or a history room, or – or whatever, I think that does often mean that art teachers do end up – much like PE [physical education] teachers, actually, as well, often do end up with disclosures being – coming through them, like, accidentally, or – or on purpose, sort of thing, because the relationship is different, because of the nature of the way that those rooms will operate, for the vast majority.” (Interview, Other-13) |
| **Art freedom** | “If I’m seeing young people who do a lot of art in school, the associations with them needing to create good art, or – I say ‘good’ in, like, in inverted commas. And I suppose I have to kind of make clear that I, I’m not there in an academic sense. I’m not there to teach them how to do something, but actually this is a space for them to use, in whichever way they want and, I suppose they’re not used to having so much freedom [laughs] in a school space.” (Interview, Creative-Arts-Therapist-2). |
|  | “As an art teacher, I find the curriculum has become preoccupied with grades, literacy and numeracy. The therapeutic nature of art has been lost within this. Allowing learners to create the art they want to, without the pressures of grading would be very beneficial to their well-being.” (Survey, Art-Teacher-10) |
|  | “I think that making art promotes positive mental health when students are able to make art with more flexibility. It is a great way for them to express and communicate without having to use words. However, due to the constraints of the curriculum/number of students in lesson/practicality of getting out different types of equipment etc, it isn't possible to do that in art lessons.” (Survey, Art-Teacher-9). |
|  | “You know, because when they go to art class in a school, you know, they’re told what to do, you know, and they compare each other, they compare their work to other’s work, you know, so there’s that separation, you know, trying to separate school art from erm, you know, from, from wellbeing art” (Interview, Counsellor-3). |
|  | “I just think that within school you're, we are sometimes, held to account for delivering within a spec or within a, you know, a scheme of learning, for example. And we're trying to hit certain things. So for me, visual art in the context of mental health and well-being is probably art that’s done for a bit more freedom if that makes sense.” (Interview, Art-Teacher-11). |
|  | “You don’t have to be a great footballer to enjoy football, you don’t have to be a great artist to enjoy drawing something, you know? You can just have a go and enjoy it” (Interview, Pastoral-Team-4). |
|  | “I think that’s another problem in schools at the moment like there’s such an emphasis on grading that kind of keeps that enjoyment out of it and there isn’t that kind of, that ability to be creative. Because they’re so set on hitting the objectives for getting a good grade so for a lot of kids by the end of GCSE they’re kind of put off art because it’s very prescribed so I think that’s been a bit of an issue” (Interview, Art-Teacher-10). |
| **Theme 2: Resources in schools are stretched** | |
| **Widening support** | “Since Covid I have noticed students are a lot more anxious.” (Survey, ELSA-1) |
|  | “Our pastoral team, like I said to you, has massively changed. We now – the team is about three times the size it was three years ago, and even then, we are worked to the bone. We don’t have capacity to take anything else on. So anything that alleviates any pressure anywhere else in the system is always going to be welcomed by us.” (Interview, Other-13) |
|  | “I think it would be a fabulous idea to bring something like that [the art and wellbeing programme] into school. Because it’s so busy – we’re so busy firefighting, not noticing the little fires that have started, and then turned into a, you know, massive fire. We’re just – we’re just leaving it all until it’s too late” (Interview, Other-4) |
|  | “The Safeguarding team, because of COVID, have been in a, an element of crisis response in terms of responding to the immediate kind of needs that are in front of us whereas we are trying to move back to a position where we are implementing things that make sure that our students remain mentally healthy throughout rather than having to just respond to cri…” (Interview, Mental-Health-Lead-3). |
|  | “I think it’s important for everybody to have access to do it, and be able to take part in it, because you never know when they might be affected by something that does affect their mental health” (Interview, Head-of-Art-8) |
| **No one in school has time** | “A school day is tightly governed by the clock, tightly governed by, ‘You’ve gotta be here, you’ve gotta be there,’ and you don’t – you don’t get kids in school with free time, and you don’t get adults with free time. Everyone, from whether you’re the kid, when you start, or the adult, when you start, to when you finish in the afternoon, it’s all – it’s completely rigid. It’s all there. Time, place, people. And, I – I would really encourage you to have that in your thinking.” (Interview, SENCO-4) |
|  | “It’s a constant, every single year. ‘You’re here to learn, you’re here to learn, you must regurgitate it.’ And there isn’t any time to fit in any – any other fun or mental health stuff.” (Interview, Other-4). |
|  | “A counsellor would make sure that their client is 'safe' to leave the session but educators would be unable to do this due to time constraints and a lack of training” (Survey, Art-Teacher-9). |
|  | “I was thinking about, was how you would actually deliver it and there’s also the problem with who could they let off timetable to do it. Because they’re very poor now state schools, a lot of them are and they’re very stretched.” (Interview, Counsellor-1). |
|  | “Usually, teachers tend to think that what they do, their subject is the most important thing ever [laughs] and the children cannot be withdrawn because it would be a waste of time” (Interview, Other-2)  “Now their attitudes towards us has completely changed [laughs]. So yeah, no because sometimes, you know, they don’t know and I think it’s very important to make people aware of this kind of mental health conditions and problems.” (Interview, Other-2). |
|  | “I feel that art teachers would be fantastic at helping with supporting this but this would need to be factored into timetabling as most teachers have a very heavy workload.” (Survey, Art-Teacher-9). |
|  | “You know part of the theme of what I’ve been saying to you is that you’re taking busy people away from something else, and to some extent. In Year 7, not such an issue, but you’re taking, you know, a group of kids out of their learning. I think that you need to have – if – if you’re looking at it being a repetitive long-term thing, I think that you need to be having a strategic meeting with, senior people in the school” (Interview, SENCO-4). |
|  | “All staff in schools are allocated roles, asking staff to step out of role to do something different causes disruption and knocks onto other staff” (Survey, SENCO-4) |
| **School staff are overworked and under-supported** | “School staff are very stressed, they’re very overworked, underpaid, mental health is not great in schools” (Interview, EMHP-3) |
|  | “I think It's important to recognise that art therapy and similar programs can be emotionally demanding, and those involved need to be prepared for this aspect of the work, so the appropriate training and supervision would be needed.” (Survey, Other-9) |
|  | “I think that as teaching staff we are so busy with day to day school life but having a external support for planning to deliver a programme and prepared resources and materials would be great” (Survey, Art-Teacher-2). |
|  | “I would say erm personally as a teacher, like it is – I do find myself – I run myself very thin.” (Interview, Art-Teacher-1) |
|  | “I wouldn’t want the group to be, like, an extra burden, or not be given the attention it needs because, like, there’s an art exam coming up, for example, and that needs to take priority, but I think the same could be said for any staff member. I think it’s an extra bit that has to be added onto their role and they need to therefore be given the time and the skills and resources to be able to do that, rather than just expect to be, to also be able to do that within their role as an art teacher.” (Interview, Creative-Arts-Therapist-2). |
|  | “I think if staff have to deliver it, and they haven’t necessarily had the chance to be part of the training, then I think they would just think it’s another add-on. And again, it’s a shame, but that is … yeah, teachers are just really stretched, so I think anything outside of what they’re already delivering feels like an add-on, unless they’re involved, and it’s made – made to have value, if that makes sense.” (Interview, Head-of-Art-8). |
|  | “As teachers, we’re humongously busy, and have a million and one other things to do, but I think that, the way society is means that actually more of us need mental health support training anyway, in schools” (Interview, Head-of-Art-1) |
|  | “On the wellbeing side I would imagine most art teachers would be able to – that’s we kind of do on a daily basis anyway really isn’t it” (Interview, Art-Teacher-10) |
|  | “You then are stepping into a different role. Rather than a role that is guiding them to create the best possible image, you’re stepping into a role that supports them in how they’re feeling, and accepting that the way they want to make imagery is the best way for them to journey through that. So it’s – like, it’s really important, I think. The number of conversations that I’ve had with students over the years about different issues that could or do affect their mental health is too many to count” (Interview, Head-of-Art-8). |
|  | “Thinking about staff’s wellbeing delivering that and that’s just from conversations I’ve had with a lot of teachers, who’ve shared they kind of feel like they’re a teacher, they’re a therapist. They’re doing… They’re social workers some days and they’re doing a lot of different roles in one.” (Interview, Creative-Arts-Therapist-1), |
|  | “I think as art teachers, we are expected that we can cope more with it sometimes. Which is great, and which is why, again, it goes, kind of, with my opinion that I think art teachers could deliver a little mini unit each year as a, kind of, standalone unit that was specific for mental health because we kind of do it anyway without much support.” (Interview, Art-Teacher-11). |
|  | “Education staff aren’t always appropriately supervised at the moment around some of the struggles that they’re experiencing. So I really struggle […], I can’t see how they would be appropriately supported, especially if something came up around the therapeutic - within that therapeutic environment, that they might struggle with emotionally and I don’t think it's up to the lead facilitator to be able to support them with that, unless they are supervision trained.” (Interview, EMHP-2). |
|  | “I wouldn’t say that we’ve had any support on it, because more of the focus at our school has been on changing behavioural patterns, and, we’ve had a focus on bullying, and things like that. So we haven’t actually looked specifically at mental health, this year, even though we’ve got more students school refusing and off for mental health reasons than ever before, so it feels a bit odd that we’re not getting more training, to be honest.” (Interview, Head-of-Art-8). |
| **Theme 3: Integration, flexibility and relationships as factors for success** | |
| **Integration into school** | “I think there’s too many things where you see an initiative come in, and I say this, sort of, having experienced it, and – where a new scheme gets introduced, with an hour’s worth of training, which is not particularly hands on, and is sort of you’re told, ‘This is what’s gonna happen,’ but you’re not told how to do it, or why you’re doing it, and then it’s like, ‘Right, it’s over to you. Go and get it done.’. I think that instantly turns people off. It does in – in all areas. Whereas where we’ve had success, um, with a lot of the changes that we’ve made over the last five, if not ten years, is it’s almost been like a slow build” (Interview, Other-13) |
|  | “I’ve seen lots of programmes like this, in the past, where someone will dip in, and then they dip out, and it’s meaningful for the time whilst it’s being done, but then it goes away, and it – it doesn’t have any real impact.” (Interview, Head-of-Art-1) |
|  | “I think that as a school we’re really guilty sometimes of doing things just to tick a box. And that sounds terrible, but it’s almost like, ‘Okay, this month is mental health day’ or something and it’s like, ‘Okay, put things into your slides.’ And I don’t ever wanna do something in my lesson that’s just ticking a box. If I’m going to do it I want to do it right and really embed it fully.” (Interview, Head-of-Art-2). |
|  | “If you get a positive evaluation for a program, the school, we're going to carry it on because a, it sounds really cynical now, it looks really good for OFSTED, because they've done this program and it's been successful, and what have you, if they didn't carry it on OFSTED, they'd be saying, ‘Well, why have you not carried it on? If it was so successful?’ So they kind of have to carry it on.” (Interview, Mental-Health-Lead-2). |
|  | “If you were offering staff training, to then run it in school, the longevity of it, in my opinion, wouldn’t be as strong, because it’s – things just get lost, and then if that member of staff left the school, it wouldn’t continue” (Interview, Pastoral-Team-9). |
|  | “If any risks or safety issues did come up within sessions, like, everyone would have a kind of safeguarding plan, or route, where they can kind of follow, and policies to follow. So, generally, that’s … you know, there’s – there’s kind of clear guidelines around that” (Interview, EMHP-6). |
|  | “Think part of the problem is you have to stop when the bell rings. That’s – that’s the problem, really. Which is the same for all counselling sessions, isn’t it? It’s – it’s kind of a finite thing.” (Interview, Other-4). |
|  | “I can highlight two or three people that it, that with the capacity they have at the moment would ensure that that happened. And then almost that long term, can they train the next group of people? And then can they train? And, and if we talked about that observing and kind of relearning from each other, that, that cycle just continues? So I think on the ground is where sustainability happens in schools.” (Interview, Mental-Health-Lead-3) |
| **Flexibility and choice** | “Within a school? It depends. Each school’s quite different. So some schools want you to have everything done the way that it’s done in their school and some schools are more open to it being done the way it’s done in your service. So historically, I worked in a secondary school and I predominantly just worked in that school. So I had, like, a school email and a lot of my things were aligned to how that school operated. So, like, my consent forms, my emails and everything needed, to some extent, to go through the school, even though it’s employed by a charity, but other schools, the way I’m working now more is that I just kind of come in.” (Interview, Creative-Arts-Therapist-1). |
|  | “I think the thing I maybe struggle with a little bit in picturing it is even at the schools I’m in, it changes so much from school to school” (Interview, Creative-Arts-Therapist-2). |
|  | “Just it being an option, and voluntary, so like, ‘You can come if you want to, but you don’t have to,’ you know, ‘It’s up to you.’ Because, a lot of the time, I think, with educational stuff, like, if they’re doing, like, lunchtime groups, or after school clubs, it’s kind of forced upon them by teachers or parents, so just being like, ‘This is up to you, if you wanna come,’ kind of shows that, you know, they have that – that kind of control, and choice over it.” (Interview, EMHP-6). |
|  | Interviewer: “What are your thoughts on art teachers facilitating an art and wellbeing programme?”  Participant: “It’s so dependent on the person. […] I think you’d probably get really different responses, just like you would from any teacher. You know, teachers are different [laughs] and some people are so set in their ways, but some people would love it. So it’s just gonna – that’s gonna be a bit of an unknown” (Interview, Art-Teacher-5) |
| **Importance of relationships** | “I think you would definitely want someone in the room that they were familiar with. Someone who is a reassuring face, a head of year or an art teacher that they’re familiar with or even just seeing the adults in the room see the value of this as well.” (Interview, Head-of-Art-2) |
|  | “I think it’s good for me, as well, like, external, because I can come in, they don’t know me, and I can build those really, like, positive relationships from the start. Whereas, of course, some children aren’t gonna get – get along with some teachers, and vice versa” (Interview, EMHP-6) |
|  | “I think it’d be great if, like, some staff members could be involved, just because they’re there at school every day, they’re familiar with the children and young people. Because, like, as me, as an external practitioner, kind of coming in, I only see them once a week. So having, like, someone there that they can go up to and be like, ‘Oh, remember when I spoke about this in, you know, our art group,’ kind of – d’you know what I mean? And, like, building that relationship up.” (Interview, EMHP-6) |
|  | “It’s quite helpful having good links with the school, because if young people need to bring forms back, or if there’s a question or something comes up, they often have a point of contact who can contact you” (Interview, Creative-Arts-Therapist-1). |
|  | “I think it needs to be someone largely that doesn’t teach, I would say. As someone that is in both right now, I still teach some lessons, but I also deal with some really difficult safeguarding concerns. I’ve noticed a shift in myself that I won’t tell a student off as much as I used to because I’m very aware that I could very much have them disclosing something, or, you know, I want them to be able to do that. So I think it is very difficult when you have somebody that’s trying to do both.” (Interview, Mental-Health-Lead-3). |
|  | “I think choosing the person to teach that has got to be done quite carefully. Because we have one art teacher in our school who many of my students don’t gel with. So, I think you’d have to make sure that they were right for delivering something.” (Interview, Other-4) |
