## Supplementary figures and images for "Visual art for the prevention of student mental health difficulties in schools: Mixed-methods analysis of staff surveys and interviews"

### S1 Figure

S1 Figure. Participant recruitment flow diagram
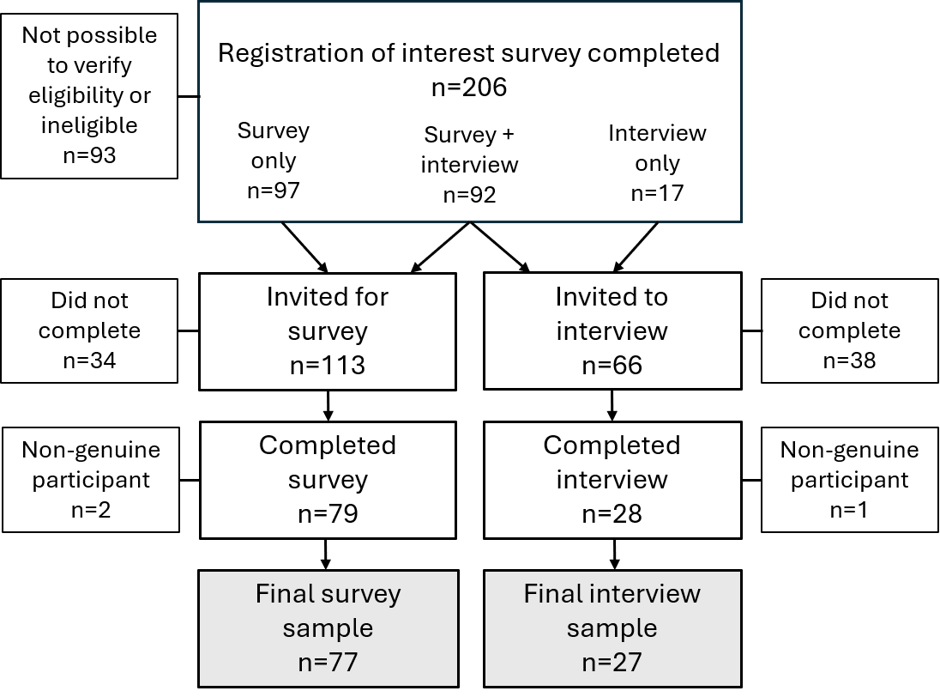
